# Effect of various treatment modalities on the novel coronavirus (nCOV-2019) infection in humans: a systematic review & meta-analysis

**DOI:** 10.1101/2020.05.24.20111799

**Authors:** Shubham Misra, Manabesh Nath, Vijay Hadda, Deepti Vibha

## Abstract

**Background and aim:** Several therapeutic agents have been investigated for the treatment of novel Coronavirus-2019 (nCOV-2019). We aimed to conduct a systematic review and meta-analysis to assess the effect of various treatment modalities in nCOV-2019 patients.

**Methods:** An extensive literature search was conducted before 22 May 2020 in PubMed, Google Scholar, Cochrane library databases. Quality assessment was performed using Newcastle Ottawa Scale. A fixed-effect model was applied if I^2^ <50%, else the results were combined using random-effect model. Risk Ratio (RR) or Standardized Mean Difference (SMD) along-with 95% Confidence Interval (95%CI) were used to pool the results. Between study heterogeneity was explored using influence and sensitivity analyses & publication bias was assessed using funnel plots. Entire statistical analysis was conducted in R version 3.6.2.

**Results:** Eighty-one studies involving 44 *in vitro* and 37 clinical studies including 8662 nCOV-2019 patients were included in the review. Lopinavir-Ritonavir compared to controls was significantly associated with shorter mean time to clinical improvement (SMD -0.32; 95%CI -0.57 to -0.06) and Remdesivir compared to placebo was significantly associated with better overall clinical improvement (RR 1.17; 95%CI 1.07 to 1.29). Hydroxychloroquine was associated with less overall clinical improvement (RR 0.88; 95%CI 0.79 to 0.98) and longer time to clinical improvement (SMD 0.64; 95%CI 0.33 to 0.94), It additionally had higher all-cause mortality (RR 1.6; 95%CI 1.26 to 2.03) and more total adverse events (RR 1.84; 95% CI 1.58 to 2.13).

**Conclusion:** Our meta-analysis suggests that except *in vitro* studies, no treatment till now has shown clear-cut benefit on nCOV-2019 patients. Lopinavir-Ritonavir and Remdesivir have shown some benefits in terms less time to clinical improvement and better overall clinical improvement. Hydroxychloroquine use has a risk of higher mortality and adverse events. Results from upcoming large clinical trials must be awaited to draw any profound conclusions.

## Introduction

The novel Coronavirus-2019 (nCOV-2019) has now encompassed more than 200 countries since a cluster of cases were initially reported in Wuhan, China on 31^st^ December 2019 (1). As of 22^nd^ May 2020, 5,322,698 people have been infected globally from nCOV-2019 while 340,319 have died of this severe infection (2), The nCOV-2019 belongs to the Coronaviridae family and has structural similarities to the betacoronavirus that has caused two epidemics in the past 18 years; Severe Acute Respiratory Syndrome coronavirus (SARS-CoV) and Middle East respiratory syndrome coronavirus (MERS-Cov) (3).

No drug or therapeutic agent has yet been approved by the United States-Food and Drug Administration (US-FDA) for treating nCOV-2019 pneumonia patients. Based on the initial results obtained from certain *in vitro* studies (4,5), non-randomized trials (6) and interim analysis of some randomized controlled trials (RCTs) (7), Hydroxychloroquine and Remdesivir received FDA emergency use authorization for nCOV-2019 (8,9). However, recent RCTs published on these respective drugs have shown inconclusive evidence for their usage in nCOV-2019 patients (10,11). Recently, FDA cautioned against the use of Chloroquine or Hydroxychloroquine outside clinical trial settings due to high risk of associated adverse events (12), With more than 500 trials already registered in clinicaltrials.gov on the treatment of nCOV-2019, it is imperative to investigate the available evidence till date and assess each treatment in terms of benefit or harm to the nCOV-2019 patients.

The aim of this systematic review and meta-analysis was to pool the initial evidence available from RCTs, non-RCTs, observational and *in vitro* studies for analyzing the benefit/harm of various treatment modalities administered to nCOV-2019 pneumonia patients. The results of this systematic review and meta-analysis might be useful in designing future clinical trials and providing guidelines.

## Methods

### Electronic search

Electronic databases including, PubMed, EMBASE, Medline, Google Scholar, Cochrane library and clinicaltrials.gov were searched independently by two authors (SM and MN) till 22 May 2020. The following MeSH terms or free text terms were used: “2019 novel coronavirus”, “2019 nCOV”, “COVID19’’, “SARS-CoV-2”, “drug therapy”, “vaccine”, “anti-viral therapy”, “symptomatic treatment”, “preventive therapy”, “immunotherapy”. The detailed search criteria are given in the supplementary file S1. Furthermore, any abstract proceedings in scientific conference and reference list of all the relevant identified articles were thoroughly searched. Grey literature and preprints were searched using the https://www.medrxiv.org and https://www.biorxiv.org databases. There were no restrictions on language. Studies published on human subjects after 31^st^ December 2019 since the nCOV-2019 outbreak initiated, were only searched. The protocol for this systematic review and meta-analysis was registered in PROSPERO (ID: CRD42020175792) and there were no major deviations from the published protocol in PROSPERO.

### Population

Subjects diagnosed with pneumonia caused by new Coronavirus 2019 infection (nCOV-2019).

### Intervention

Various treatment modalities for nCOV-2019 patients.

### Comparator

nCOV-2019 patients receiving standard care or placebo treatment or secondary treatment drug.

### Outcome

#### Outcome for *in vitro* studies

Inhibition potential and Cytotoxicity of the drug

#### Outcome for clinical studies

(1) All-cause Mortality; (2) total adverse events; (3) overall clinical improvement defined as the number of patients becoming negative for nCOV-2019 or improvement in overall symptoms or discharged from the hospital; (4) time to clinical improvement defined as the time in number of days from treatment initiation to becoming negative for nCOV-2019 or time in number of days from treatment initiation to improvement in overall symptoms or time in number of days from treatment initiation to discharge from the hospital.

### Inclusion and exclusion criteria

The criteria for the inclusion of studies in our systematic review and meta-analysis was: For inclusion of clinical studies: (1) randomized controlled trials (RCTs), non-RCTs, cohort studies, case-control studies; (2) studies should be focused on various treatments given to nCOV-2019 patients; (3) studies must have a comparator group comparing the primary treatment drug to either standard care/control or placebo or a second treatment drug; (4) conducted on human subjects only.

For inclusion of *in vitro* studies: (1) case series, observational studies; (2) studies should be focused on various treatments given for nCOV-2019, (3) studies should have reported data on inhibitory effect and cytotoxicity of the drug.

The following clinical and *in vitro* studies were excluded from our systematic review and meta-analysis: (1) conducted on animal models; (2) completed studies but results not published or preprints not available; (3) ongoing registered clinical trials; (4) desired outcome data not reported; (5) single arm studies/trials (for clinical studies). This systematic review and meta-analysis was performed according to the Preferred Reporting Items for Systematic Reviews and Meta-Analyses Protocol (PRISMA-P) 2015 guidelines (13).

### Data extraction

All titles and abstracts retrieved by searching available literature were screened independently by two authors (SM and MN) against the eligibility criteria. The information extracted from each eligible study included the first author, year of publication, study design, sample size, interventions (including type of treatment administered), outcome measures, main results. Any disagreement was resolved by mutual consensus among all the authors.

### Risk of bias assessment

The quality assessment was done for only the clinical studies included in our systematic review and meta-analysis by two independent authors (SM and MN) using the Newcastle-Ottawa Scale (14). Since we pooled the results from both RCTs as well as non-RCTs and cohort studies, the quality assessment of RCTs was done considering them as cohort studies only. Any disagreement was resolved by consulting with the remaining authors of the review.

The risk of publication bias was assessed by using Funnel plots and the asymmetry of the funnel plot was investigated using the Egger’s regression test (15).

### Statistical analysis

Dichotomous variables were represented by number (percentage) and the continuous variables were represented by mean and standard deviation (SD), If median, ranges and/or interquartile range were reported, then they were converted to mean and SD using the formula depending upon the sample size given by Wan et al. 2014 (16). A meta-analysis was performed only for clinical studies and for those treatments in which required outcome data could be pooled from two or more studies. For dichotomous variables, the data was pooled using Risk ratio (RR) and 95% Confidence Interval (95% CI) while for continuous variables, the data was pooled using Standardized Mean Difference (SMD) and 95%CI. Heterogeneity among the included studies was investigated using Cochran’s Q statistic, I^2^ metric tests and by using prediction intervals. A fixed-effect model was applied if I^2^ was less than 50%, else a random-effect model was used to pool the results. Labbé plots were used to determine the trend and between-study heterogeneity present in a binary outcome meta-analysis. The source of heterogeneity was further assessed by using the influence diagnostic tools and by conducting the sensitivity analyses and meta-regression analyses. All the statistical analysis was conducted using R version 3.6.2.

## Results

Initial search yielded 1479 articles by searching various databases for published and preprint articles. After screening 917 articles, 355 full text articles were reviewed for eligibility and finally 81 studies were included in our systematic review and meta-analysis. Further, out of the 81 included studies, 44 were *in vitro* studies and 37 were clinical studies. The meta-analysis was finally conducted on 21 clinical studies. Fig 1 represents the PRISMA flow diagram for the inclusion of studies in our systematic review and meta-analysis.

**Figure 1:**
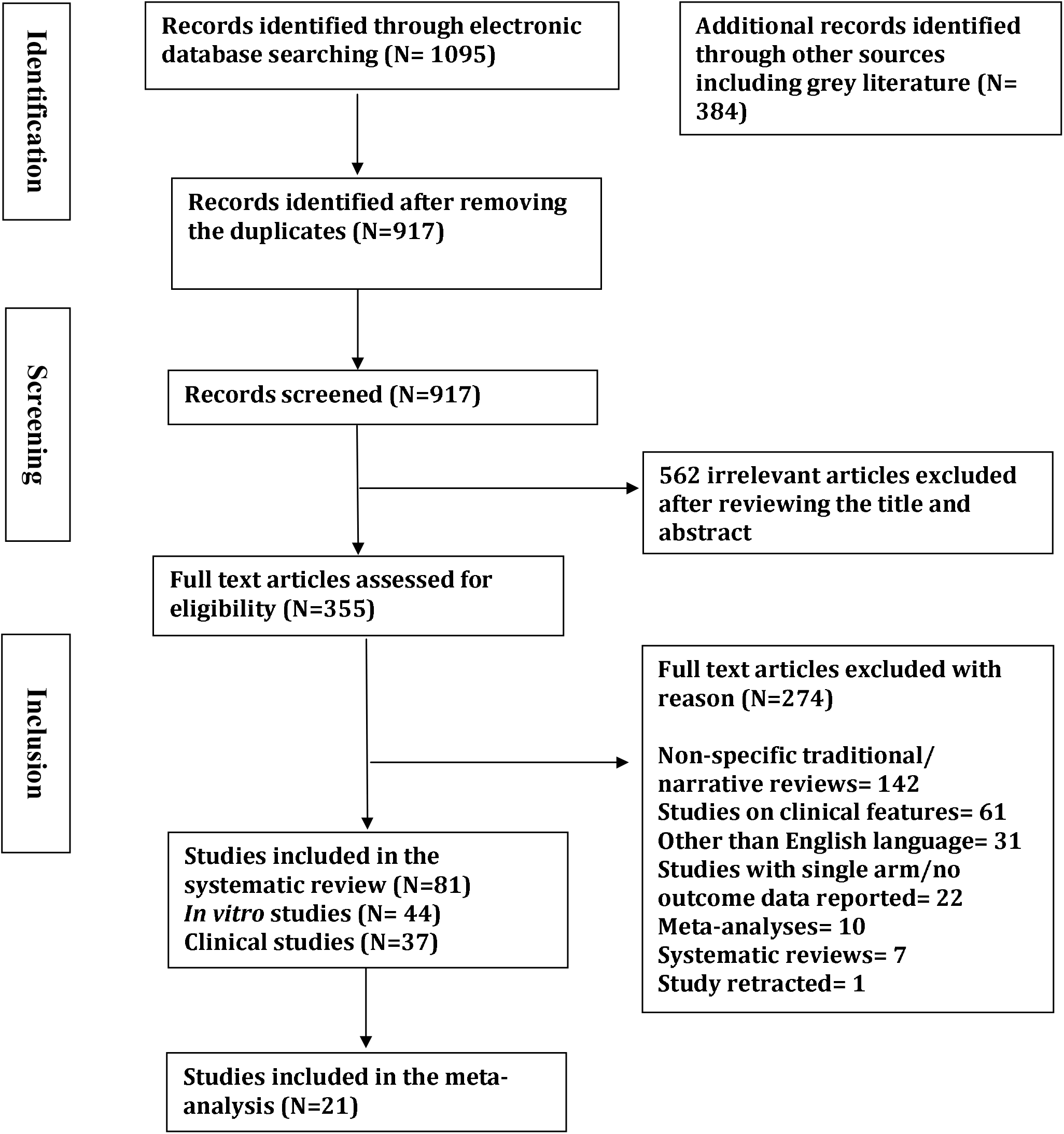
PRISMA flow diagram for the systematic review and meta-analysis.

### Results from the systematic review of *in vitro* studies

Overall, 44 *in vitro* studies were included in the systematic review, which comprised majorly of treatments done on Vero E6 cells for viral titration, drug inhibition and cytotoxicity analyses. There were 32 preprints and 12 published articles amongst the selected studies. Fifteen studies were included from China(17-31), six from United States of America (USA) (32-37), five from France(38-42), four from Japan(43-46), two each from Germany(47,48), Netherlands(49,50) and South Korea(51,52) and one each from Australia(53), Brazil(54), Canada(55), Israel(56), Italy(57), Norway(58), Switzerland(59) and the United Kingdom (UK) (60).

There were five studies(23,30,37,59,60) involving the anti-malarial drug Chloroquine with multiplicity of infection (MOI) ranging from 0.01 to 0.05 except one that did not report any MOI, had average half-maximal inhibitory concentration (IC50) of 25.94 μM [1.03 - 46.8], average cytotoxic concentration (CC_50_) of 40.48 μM [16.76 - 50] and average selectivity index (SI) of 34.92 [1.19 − 88.5]. All the studies used Vero E6 cells for anti-nCOV-2019 inhibition with Chloroquine treatment *in vitro* and reported significant inhibitory effect on viral population despite having some form of cellular cytotoxicity.

Six studies(30,31,36,37,41,52) involving the broad-spectrum antiviral Remdesivir with MOI in the range of 0.01 to 0.1 exhibited mean half-maximal effective concentration (EC_50_) of 6.7±0.085(μM [0.72−23.15], average IC50 of 1.9μM [1.3-2.5], average CC_50_ of 137.5 (μM (100−175) and average SI of 99.935 (70−129.87), Remdesivir showed promising results *in vitro* and depicted potency inhibiting viral proliferation particularly in Vero E6 and Calu-3 cell lines.

There were four studies(21−23,38) on Hydroxychloroquine out of which three had positive controls (two studies with chloroquine and one with Remdesivir), The studies had a wide range of MOI from 0.002 to 0.8 and an average EC_50_ of 5.95μM [0.72 − 4.17] in the treatment arm as compared to 4.83μM [1.65−7.36] in the control arm. The average CC_50_ of 101.58μM [15.26−249.5] and average SI of 23.8 (10−61.45) resulted in a positive outcome in all the four studies where hydroxychloroquine showed better efficacy and lesser cytotoxicity in inhibiting nCOV-2019 than Chloroquine *in vitro*.

Three studies(45,46,58) on the anti-retroviral drug Nelfinavir (MOI: 0.01−0.1) had an average EC_50_ of 1.61±0.6μM [1.13−2.1±0.6], CC_50_ of 32.67 ± 0.4 μM [9.7±0.4−64] and average SI of 13.6 [4.6−21.52], Nelfinavir had been found to be highly potent when administered in combinatorial approach as well as stand-alone. While, two studies(31,44) on Lopinavir (MOI: 0.01−0.02) depicted an average EC_50_ of 16.18μM [5.73-26.63] and average CC_50_ of 62.09μM [49.75−74.44] which resulted in lesser efficacy less than Nelfinavir and Remdesivir *in vitro*.

Two studies each of Chlorpromazine(39,60), Ciclesonide(43,51), Protoporphyrin IX(27,28) and Nafamostat mesylate(44,52) highlighting their inhibitory efficacy depicted that Chlorpromazine (EC_50_ = 9μM, IC50 = 4.03μM), Ciclesonide (EC_50_ = 4.4μM, IC50 = 4.33μM) and Protoporphyrin IX (EC_50_ = 0.23μM, IC50 = 0.11±0.02μM) effectively inhibited nCOV-2019 infected cell lines. However, Nafamostat mesylate (EC_50_ = 0.0115μM, IC_50_ = 0.0022μM) was found to be one of the most potent inhibitors of nCoV-2019 infection *in vitro*(44,52).

One study each involving 47D11 H2L2 antibody, Arbidol(17), Atazanavir(54), Auranofin(32), Baicalein(18), Beta-d-N4-hydroxycytidine(33), Boceprevir(34), Darunavir(47), Genz-123346 (GlucosylCeramide synthase inhibitor)(56), Antibodies n3086/n3113(20), Interferon-α/ Interferon −(β(35), Indomethacin(24), Lianhuaqingwen(25), Miglustat(57), Naproxen(40), Pudilan Xiaoyan Oral Liquid(29), Suramin(50) and T-705 (Favipiravir)(42) had a wide range of EC_50_ [0.15−207 μM] or IC50 [0.08−411.2μM] values. However, each of these compounds demonstrated the potential to stall the process of viral replication and growth through inhibiting viral titre in cell lines.

Two studies(34,55) of the novel protease inhibitor GC376 depicted an average EC_50_ of 2.14±1.68μM and average CC_50_> 150μM suggests that these novel inhibitors of nCOV-2019 infected Vero E6 cells have the potential to be up-scaled into further animal model studies and clinical trials.

There was one *in vitro* study(19) on the phytochemical extracts from six Chinese traditional medicinal plants viz. *Cimicifuga rhizoma, Meliae cortex, Coptidis rhizoma, Phellodendron cortex, Sophora subprostrata radix* and *Mountan cortex radicis*. Extracts from five of the six plants showed potential as herbal medicine by inhibiting coronavirus infection in both A59 and Vero cells with significant EC_50_ [2.0±0.5−27.5±1.1μg/mL], CC_50_ [71.3 ± 7.2−334.3 ± 7.0μg/mL] and SI [11.1−34.9] values.

All the studies had an incubation time (hours post infection) of treatment ranging from 24 to 72 hours on average with a median of 48 hours. Table-1 depicts the baseline characteristics of *in vitro* studies included in our systematic review.

**Table 1:** Baseline characteristics of *in vitro studies* on the inhibition and potential treatment of nCOV-2019.

| S. No. | Author, Year | Country | Sample type | Assay Used | Treatment | Control | EC<sub>50</sub> or IC<sub>50</sub> | Incubation time (h) | Publication Status |
| --- | --- | --- | --- | --- | --- | --- | --- | --- | --- |
| 1 | Wang C, 2020(49) | Netherlands | Vero E6 | VNA | 47D11 H2L2 antibody | Isotype | 0.15ug/ml | 24 | Published |
| 2 | Wang X, 2020(17) | China | Vero E6 | CCK8 | Arbidol | - | 4.11 [3.55-4.73] uM | 48 | Published |
| 3 | Fintelman-Rodrigues N, 2020(54) | Brazil | Vero E6; A549 | CPE; RT-PCR | ATV; ATV/RTV) | CQ | Vero: ATV/RTV = $0.5 \pm 0.08 \mu\text{M}$; ATV = $2.0 \pm 0.12 \mu\text{M}$; A549: ATV/RTV = $0.60 \pm 0.05 \mu\text{M}$; ATV = $0.22 \pm 0.02 \mu\text{M}$ | 48 | Preprint |
| 4 | Rothan H, 2020(32) | USA | Huh7 | q-RT-PCR | Auranofin | DMSO | 1.4uM | 24, 48 | Preprint |
| 5 | Touret F, 2020(38) | France | Vero E6 | CPE | AZM, HCQ, ODH, QHM, APS | RDV | AZM = 2.12, HCQ = 4.17, ODH = 5.05, QHM = 5.11, APS = 5.39 uM | 48 | Preprint |
| 6 | Su H, 2020(18) | China | Vero E6 | CCK8; qRT-PCR | Bl; Ble | DMSO | Bl = 10.27uM, Ble = 1.69uM | 48 | Preprint |
| 7 | Sheahan T, 2020(33) | USA | Calu 3, Vero E6 | CTG; Plaque | NHC | - | Vero: 0.3, Calu3: 0.08uM | 48, 72 | Published |
| 8 | Ma C, 2020(34) | USA | A549, MDCK, HCT-8, Caco-2, Vero, and BEAS2B | CPE | Bcr, GC-376, CII, CXII | - | Bcr = 1.90, CII = $2.07 \pm 0.76$, CXII = $0.49 \pm 0.18$, GC-376 = $3.37 \pm 1.68 \mu\text{M}$ | 48 | Preprint |
| 9 | Weston S, 2020(60) | UK | Vero E6 | CTG | CQ, CPZ | - | CQ = 42.03, 46.8; CPZ = 3.14, 4.03 | 72 | Preprint |
| 10 | Plaze M, 2020(39) | France | Vero E6, A549-ACE2 | Plaque | CPZ | - | Vero: 9uM, A549: 10.4uM | 48 | Preprint |
| 11 | Matsuyama S, 2020(43) | Japan | Vero E6 | 96-well | Ciclesonide | - | 4.4uM | 27 | Preprint |
| 12 | Kim HY, 2020(19) | China | MHV-A59; Vero | MTT | Clrh; MEco; COrh; PHco; Ssr; Mcr | No extract | Clrh = $19.4 \pm 7.0$, MEco = $13.0 \pm 1.4$, COrh = $2.0 \pm 0.5$, PHco = $10.4 \pm 2.2$, Ssr = $27.5 \pm 1.1$, Mcr = $61.9 \pm 6.1 \mu\text{g/ml}$ | 12 | Published |
| 13 | Meyer S, 2020(47) | Germany, USA, Belgium | Caco-2 | MTT; CPE | Darunavir | RDV | >100uM | 48 | Preprint |
| 14 | Vuong W, 2020(55) | Canada | Vero E6, A549 | Plaque | GC373 and GC376 | - | GC373 = $1.5 \mu\text{M}$, GC376 = $0.92 \mu\text{M}$ | 48 | Preprint |
| 15 | Vitner E, 2020(56) | Israel | Vero E6 | LDH | Genz-123346, GENZ-667161 | - | GZ-161 = GZ-346 = 2uM | 48 | Preprint |
| 16 | Wu Y, 2020(20) | China | Vero E6 | CPE | Group E antibodies: n3086, n3113 | - | n3086 = 26.6, n3113 = 18.9 ug/ml | 3 | Preprint |
| 17 | Yao X, 2020(21) | China | Vero | 96 Well | HCQ | CQ | 0.72uM | 24, 48 | Published |
| 18 | Liu J, 2020(22) | China | Vero E6 | CCK8 | HCQ | CQ | 12.96uM | 48 | Published |
| 19 | Yang J, 2020(23) | China | H9C2, Hep3B, HEK293, Vero, IMR-90, A549, ARPE-19, IEC-6 | CPE | HCQ, CQ | - | CQ/HCQ: H9C2 = 17.1/15.26, HEK293 = 16.76/15.26 uM (CC<sub>50</sub>) | 48, 72 | Preprint |
| 20 | Mantlo E, 2020(35) | USA | Vero E6 | CPE | IFN-α, IFN-β | - | IFN-α = 1.35, IFN-β = 0.76 IU/ml | 22 | Published |
| 21 | Xu T, 2020(24) | China | Vero E6 | 96 Well; MTT | Indomethacin | - | 1uM | 48 | Preprint |
| 22 | Caly L, 2020(53) | Australia | Vero/hSLAM | 12-well; Taqman RT-PCR | Ivermectin | Viral DNA | 2uM | 0, 72 | Published |
| 23 | Runfeng L, 2020(25) | China | Vero E6 | MTT; CPE; Plaque | Lianhuaqingwen (LH) | RDV | 411.2 μg/mL | 72 | Published |
| 24 | Rajasekharan S, 2020(57) | Italy | Vero E6 | Plaque; Cytotoxicity | Miglustat | - | 41±22 μM | 72 | Preprint |
| 25 | Yamamoto M, 2020(44) | Japan | Calu-3; VeroE6 | CPE | NM | - | Pre - 0.0115uM; No_pre - 3.16 | 72 or 120 | Preprint |
| 26 | Ko M, 2020(52) | South Korea | Calu3 | Plaque | NM, RDV, Dt | - | NM = 0.0022, RDV = 1.3, Dt = 0.16 uM | 24 | Preprint |
| 27 | Terrier O, 2020(40) | France | Vero E6 | RMS; qRT-PCR | Naproxen | No drug | 59.53uM | 48, 72 | Preprint |
| 28 | Yamamoto N, 2020(45) | Japan | Vero E6 | 96 well; qRT-PCR | NFV, LV, RTV | - | NFV = 1.13, LV = 5.73, RTV = 8.63uM | 24 | Preprint |
| 29 | Ohashi H, 2020(46) | Japan | Vero E6 | CPE | NFV, CEP | - | NFV = 0.77, CEP = 0.35 uM | 48 | Preprint |
| 30 | Jeon S, 2020(51) | South Korea | Vero | CPE | NS, CS | CQ, LV | NS = 0.28 uM; CS = 4.33uM | 72 | Preprint |
| 31 | Zheng F, 2020(26) | China | Vero E6 | Plaque | Novaferon | - | 1.02ng/ml | 48 | Preprint |
| 32 | Gu C, 2020(27) | China | Vero E6 | CCK8; 96 well; qRT-PCR | PPIX, VP | RDV | P IX = 0.23; VP = 0.03 μM | 48 | Preprint |
| 33 | Lu S, 2020(28) | China | Vero E6 | MTT | PPIX | No drug | Pre-treat = 0.21±0.02; Full-time treat = | 24 | Preprint |
|  |  |  |  |  |  |  | 0.11±0.02 µM |  |  |
| 34 | Deng W, 2020(29) | China | Vero E6 | CPE | PDL | - | 1.078 mg/mL | - | Published |
| 35 | Wang M, 2020(30) | China | Vero E6 | CCK8 | RDV, CQ | NA | RDV- 0.77uM, CQ - 1.13uM | 48 | Published |
| 36 | Choy KT, 2020(31) | China | Vero E6 | TCID50; qRT-PCR; CTG; LCVA | RDV, LV, EH, HH | - | RDV = 23.15uM, LPV = 26.63, EH = 0.46, HH = 2.55 | 48 | Published |
| 37 | Pruijssers A, 2020(36) | USA | Vero E6, Calu3 2B4 | CTG; CVA; qRT-PCR | RDV, GS-441524 | - | Calu3: RDV(2.17±0.14 µM); GS(0.85±0.16 µM) | 24, 48, 72 | Preprint |
| 38 | Pizzorno A, 2020(41) | France | Vero E6 | RMS; qRT-PCR | RDV, RDV-DZ | - | RDV= 0.72±0.03; RDV+DZ = 0.35±1.20uM | 48 to 72 | Preprint |
| 39 | Liu S, 2020(37) | USA | Vero E6 | RMTCID | RDV, CQ | - | RDV = 2.5uM; CQ (pre/post): 30/>50uM | 48 | Preprint |
| 40 | Holwerda M, 2020(59) | Switzerland | Vero E6 | GCA; CTG | Retro-2.1, NNDNJ, PBDNJ0803, CQ, URM-099-C | RDV | Retro-2.1 = 0.4909, CQ = 1.03, NNDNJ = 1.677, PBDNJ0803 = 2.026, URM-099-C = 0.335 uM | 24 | Preprint |
| 41 | Ianevski A, 2020(58) | Norway | Vero E6 | CTG | SLM, OCX, ADQ, NFV, EH, and HH | No drug/ Mock treatment | EMT = <0.01, HH= 0.03±0.02, OCX = 0.3±0.1, SLM = 0.2±0.1, ADQ = 4.2±0.2, NFV = 2.1±0.6 uM | 72 | Preprint |
| 42 | Gassen N, 2020(48) | Germany | VeroFM | Plaque | SPD, NS, MK-2206 | - | SPD = 193 µM, MK-2206 = 0.09 µM, NS = 0.17 µM | 24, 48 | Preprint |
| 43 | Silva C, 2020(50) | Netherlands | Vero E6, Calu3 | MTT | Suramin |  | 20 ± 2.7 µM | 3 | Preprint |
| 44 | Shannon A, 2020(42) | France | Vero E6 | CPE; CTB | T-705 (Favipiravir) | - | 207.1uM | 3 | Preprint |

**Abbreviations:** Virus Neutralization assay (VNA); Cell counting kit 8 (CCK8); CellTiter-Glo (CTG); Methyl Thiazolyl Tetrazolium (MTT); Cytopathic effect (CPE) inhibition assay; Reed & Muench statistical method (RMS); Luminescent Cell Viability Assay (LCVA); Cell Viability Assay (CVA); Qualitative Reverse Transcriptase Polymerase Chain Reaction (qRT-PCR); Reed & Muench Tissue Culture Infectious Dose 50 Assay (RMTCID); Green cytotoxicity assay (GCA); CellTiter-Blue (CTB); Atazanavir (ATV); Ritonavir (RTV); Chloroquine (CQ); Remdesivir (RDV); Baicalin (Bl); Baicalein (Ble); Beta-d-N4-hydroxycytidine (NHC); Boceprevir (Bcr); Calpain inhibitors II, and XII (CII and CXII); Chlorpromazine (CPZ); Cimicifuga rhizome (Clrh), Meliae cortex (MEco), Coptidis rhizome (Corh), Phellodendron cortex (PHco), Sophora subprostrata radix (Ssr), Mountain cortex radices (Mcr); Hydroxychloroquine (HCQ); Nafamostat mesylate (NM); Digitoxin (Dt); Nelfinavir (NFV); Cepharranthine (CEP); Lopinavir (LPV); Ritnovir (RTV); Niclosamide (NS); Ciclesonide (CS); Protoporphyrin IX (PPIX); Verteporfin (VP); Pudilan Xiaoyan Oral Liquid (PDL); Ementine Hydrochloride (EH); Homoharringtonine (HH); Diltiazem (DZ); N-Nonyldeoxynojirimycin (NN-DNJ); Salinomycin (SLM), obatoclax (OCX), amodiaquine (ADQ); Spermidine (SPD); Azithromycine (AZM); Opipramol dihydrochloride (ODH); Quinidine hydrochloride monohydrate (QHM); Alprostadiol (APS); Interferon-α/ Interferon-β (IFN α/ β)

### Results from clinical studies

A total of 37 clinical studies with 8662 nCOV-2019 pneumonia patients were included in the systematic review out of which 5222 [60.29%] patients were male. The overall mean age of the subjects present in the included studies was 53.82 ± 14.63. We included 14 RCTs, 3 non-RCTs and 20 observational (including retrospective/prospective cohort, case-control) studies. Among the included studies, there were 20 published articles and 17 articles with their preprints available. Twenty-five studies were included from China (10,11,61–83), six from USA (84–89), two from France (6,90) and one each from United Arab Emirates (UAE) (91), Brazil (92), Italy (93) and Hong Kong (94).

There was no significant difference in the mean age and male gender between any of the treatment and comparator groups included in our meta-analysis. Table-2 depicts the baseline characteristics of clinical studies included in the systematic review and meta-analysis. Since only limited clinical trials and observational studies have been published till date, the data from several studies could not be pooled together to assess any of the four outcome measures. Fig S2.1 in the supplementary file S2 illustrates the effect of various treatment modalities in individual studies in terms of all-cause mortality, total adverse events, overall clinical improvement and time to clinical improvement

**Table 2:** Baseline characteristics of clinical studies included in the systematic review and meta-analysis.

| S. No | Author, Year | Country | Single/Multicentre | Study design | Total sample size | Overall age, Mean (SD) | Total males, n(%) | Treatment | Comparator | Published/Preprint | Quality score |
| --- | --- | --- | --- | --- | --- | --- | --- | --- | --- | --- | --- |
| 1 | Tang W, 2020 (10) | China | Multicentre | Phase-4, Open label RCT | 150 | 46.1 (14.7) | 82 | HCQ | Control | Published | 7 |
| 2 | Chen Z, 2020 (62) | China | Single | Phase-4, Double blind RCT | 62 | 44.7 (15.3) | 29 | HCQ | Control | Preprint | 6 |
| 3 | Chen J, 2020 (61) | China | Single | Phase-3, Open label RCT | 30 | 48.6 (3.7) | 21 | HCQ | Control | Published | 6 |
| 4 | Gautret P, 2020 (6) | France | Multicentre | Open label non-RCT | 36 | 45.1 (22) | 15 | HCQ | Control | Published | 4 |
| 5 | Geleris J, 2020 (84) | USA | Single | Observational study | 1376 | . | 781 | HCQ | Control | Published | 7 |
| 6 | Yu B, 2020 (64) | China | Single | Retrospective cohort study | 568 | 66.67 (7.43) | 358 | HCQ | Control | Preprint | 5 |
| 7 | Magagnoli J, 2020 (85) | USA | Single | Retrospective cohort study | 368 | 67.82 (11.57) | 368 | HCQ, HCQ + Azithromycin | Control | Preprint | 5 |
| 8 | Mahévas M, 2020 (90) | France | Multicentre | Retrospective observational study | 181 | 60 (11.96) | 128 | HCQ | Control | Preprint | 6 |
| 9 | Mallat J, 2020 (91) | UAE | Single | Retrospective observational study | 34 | 38.67 (13.16) | 25 | HCQ | Control | Preprint | 7 |
| 10 | Rosenberg ES, 2020 (86) | USA | Multicentre | Retrospective cohort study | 1438 | 63 | 858 | HCQ | Control | Published | 7 |
| 11 | Borba | Brazil | Single | Phase-2b, | 81 | 51.1 | 61 | Low dose CQ | High dose CQ | Published | 8 |
|  | MGS,<br>2020 (92) |  |  | Parallel,<br>double-blind<br>RCT |  | (13.9) |  |  |  |  |  |
| 12 | Huang M,<br>2020 (a)<br>(65) | China | Multicentre | Prospective<br>Observational<br>study | 373 | 44.65<br>(13.29) | 175 | CQ | Control | Preprint | 6 |
| 13 | Huang M,<br>2020 (b)<br>(66) | China | Single | Open label<br>RCT | 22 | 46<br>(16.64) | 13 | CQ | Lopinavir-Ritonavir | Published | 6 |
| 14 | Cao B,<br>2020 (67) | China | Single | Open label<br>RCT | 199 | 58.33<br>(14.19) | 120 | Lopinavir-Ritonavir | Control | Published | 7 |
| 15 | Ye XT,<br>2020 (68) | China | Single | Retrospective<br>observational<br>study | 47 | . | 22 | Lopinavir-Ritonavir | Control | Published | 5 |
| 16 | Jun C,<br>2020 (69) | China | Single | Retrospective<br>observational<br>study | 134 | 48.25<br>(5.19) | 69 | Lopinavir-Ritonavir +<br>Arbidol | Control | Published | 7 |
| 17 | Li Y, 2020<br>(70) | China | Single | Phase-4,<br>Open label<br>RCT | 86 | 49.4<br>(14.7) | 24 | Lopinavir-Ritonavir +<br>Arbidol | Control | Preprint | 7 |
| 18 | Wang Y,<br>2020 (11) | China | Multicentre | Phase-3,<br>Double-blind,<br>Placebo-controlled<br>RCT | 236 | 64<br>(11.19) | 140 | Remdesivir | Placebo | Published | 9 |
| 19 | Beigel JH.<br>2020 (89) | USA | Multicentre | Phase-3,<br>Double-blind,<br>Placebo-controlled<br>RCT | 1059 | 58.9<br>(15) | 684 | Remdesivir | Placebo | Published | 8 |
| 20 | Chen C,<br>2020 (63) | China | Multicentre | Open label<br>RCT | 236 | . | 110 | Favipiravir | Arbidol | Preprint | 6 |
| 21 | Cai Q,<br>2020 (71) | China | Single | Open label<br>non-RCT | 80 | 47.92<br>(19.06) | 35 | Favipiravir | Lopinavir-Ritonavir | Published | 6 |
| 22 | Lou Y, 2020 (72) | China | Single | Open label RCT | 29 | 52.5 (12.5) | 21 | Favipiravir, Baloxavir | Control | Preprint | 5 |
| 23 | Cantini F, 2020 (93) | Italy | Single | Open label non-RCT | 24 | 63.57 (11.95) | 20 | Baricitinib | Control | Published | 7 |
| 24 | Hung IFN, 2020 (94) | Hong Kong | Multicentre | Phase-2, Open label RCT | 127 | 48.67 (22.5) | 68 | Lopinavir/ Ritonavir, Ribavirin, IFN- B1 | Lopinavir- Ritonavir | Published | 8 |
| 25 | Deng L, 2020 (73) | China | Single | Retrospective cohort study | 33 | 44.56 (15.73) | 17 | Lopinavir/ Ritonavir, Arbidol | Lopinavir- Ritonavir | Published | 6 |
| 26 | Lan X, 2020 (74) | China | Single | Pilot retrospective study | 73 | 55.65 (14.77) | 37 | Lopinavir/ Ritonavir, Arbidol | Lopinavir- Ritonavir | Preprint | 5 |
| 27 | Carlucci PM, 2020 (87) | USA | Multicentre | Retrospective observational study | 932 | 62.43 (15.62) | 584 | Zinc sulphate, HCQ, Azithromycin | HCQ, Azithromycin | Preprint | 5 |
| 28 | Fadel R, 2020 (88) | USA | Multicentre | Quasi Experimental study | 213 | 61.96 (16.07) | 109 | Pre- corticosteroid group | Post- corticosteroid group | Published | 6 |
| 29 | Wang D, 2020 (75) | China | Single | Retrospective cohort study | 115 | 55.33 (20.27) | 58 | Corticosteroid | Control | Preprint | 5 |
| 30 | Wang Y, 2020 (76) | China | Single | Retrospective cohort study | 46 | 55.33 (12.24) | 26 | Corticosteroid | Control | Published | 6 |
| 31 | Zha L, 2020 (77) | China | Multicentre | Retrospective cohort study | 31 | 41.67 (17.1) | 20 | Corticosteroid | Control | Published | 5 |
| 32 | Lu X, 2020 (78) | China | Single | Case-Control study | 62 | 58.66 (13.6) | 32 | Corticosteroid | Control | Preprint | 8 |
| 33 | Qin N, 2020 (79) | China | Single | Retrospective observational study | 72 | 49.33 (15.89) | 41 | Corticosteroid | Control | Published | 5 |
| 34 | Shi C, 2020 (80) | China | Single | Retrospective cohort study | 42 | 65.83 (36.97) | 27 | Low Molecular Weight | Control | Preprint | 6 |
|  |  |  |  |  |  |  |  | Heparin |  |  |  |
| 35 | Zhong M,<br>2020 (81) | China | Single | Single blind,<br>placebo<br>controlled<br>RCT | 17 | 62.67<br>(5.66) | 13 | Alpha Lipoic<br>Acid | Placebo | Preprint | 8 |
| 36 | Bian H,<br>2020 (82) | China | Single | Phase-1/2,<br>Open label<br>RCT | 28 | 56.58<br>(16.83) | 16 | Meplazumab | Control | Preprint | 6 |
| 37 | Liu X,<br>2020 (83) | China | Single | Retrospective<br>cohort study | 22 | 55.27<br>(12.27) | 15 | Dipyridamole | Control | Preprint | 6 |

**Abbreviations:** HCQ- Hydroxychloroquine; CQ- Chloroquine; IFN- Interferon; USA- United States of America; UAE- United Arab Emirates

### Results from the meta-analyses of clinical studies

#### Hydroxychloroquine Versus Control groups

Ten studies consisting of 3184 nCOV-2019 cases were included in the meta-analysis and were divided into two groups: 1473 subjects to Hydroxychloroquine group and 1711 subjects to control group. Compared to the control group, Hydroxychloroquine was neither found to be significantly associated with an increase or decrease in the all-cause mortality (RR 1.22; 95%CI 0.72 to 2.07) nor was found to be significantly associated with overall clinical improvement (RR 0.92; 95%CI 0.82 to 1.04) and time to clinical improvement (SMD 0.33; 95%CI -0.28 to 0.93). Although, through the Labbé plots, we did observe a trend that all-cause mortality was more towards the Hydroxychloroquine group (supplementary file S2) while overall clinical improvement was more in the control group (Fig 8(a)). However, Hydroxychloroquine had an increased risk of having total adverse events compared to the control group (RR 1.84; 95% CI 1.58 to 2.13) (Fig 2(a-d)).

**Figure 2(a-d):**
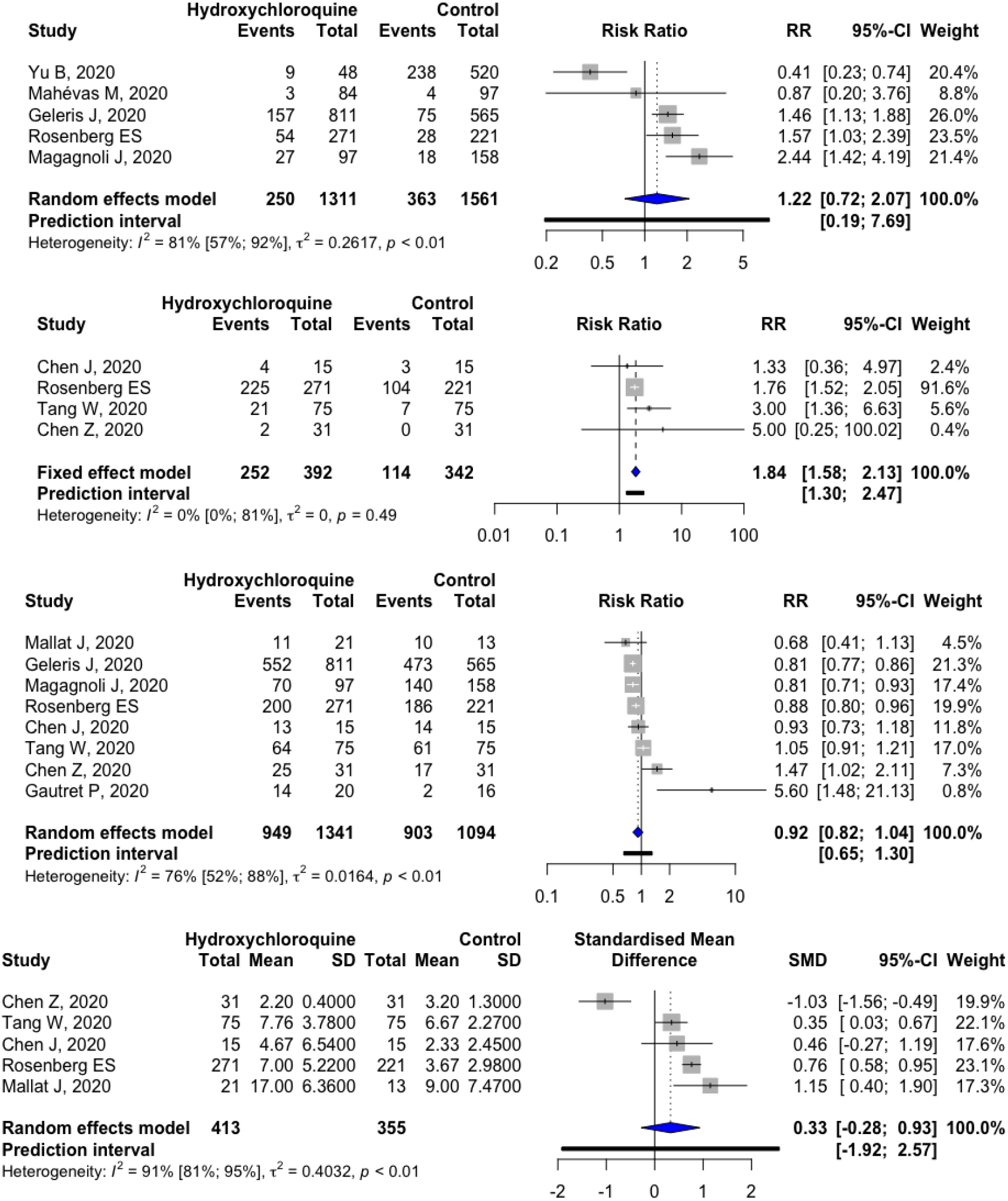
Meta-analysis of hydroxychloroquine vs. control groups to assess (a) all-cause mortality, (b) total adverse events, (c) overall clinical improvement, (d) time to clinical improvement.

**Figure 3(a-c):**
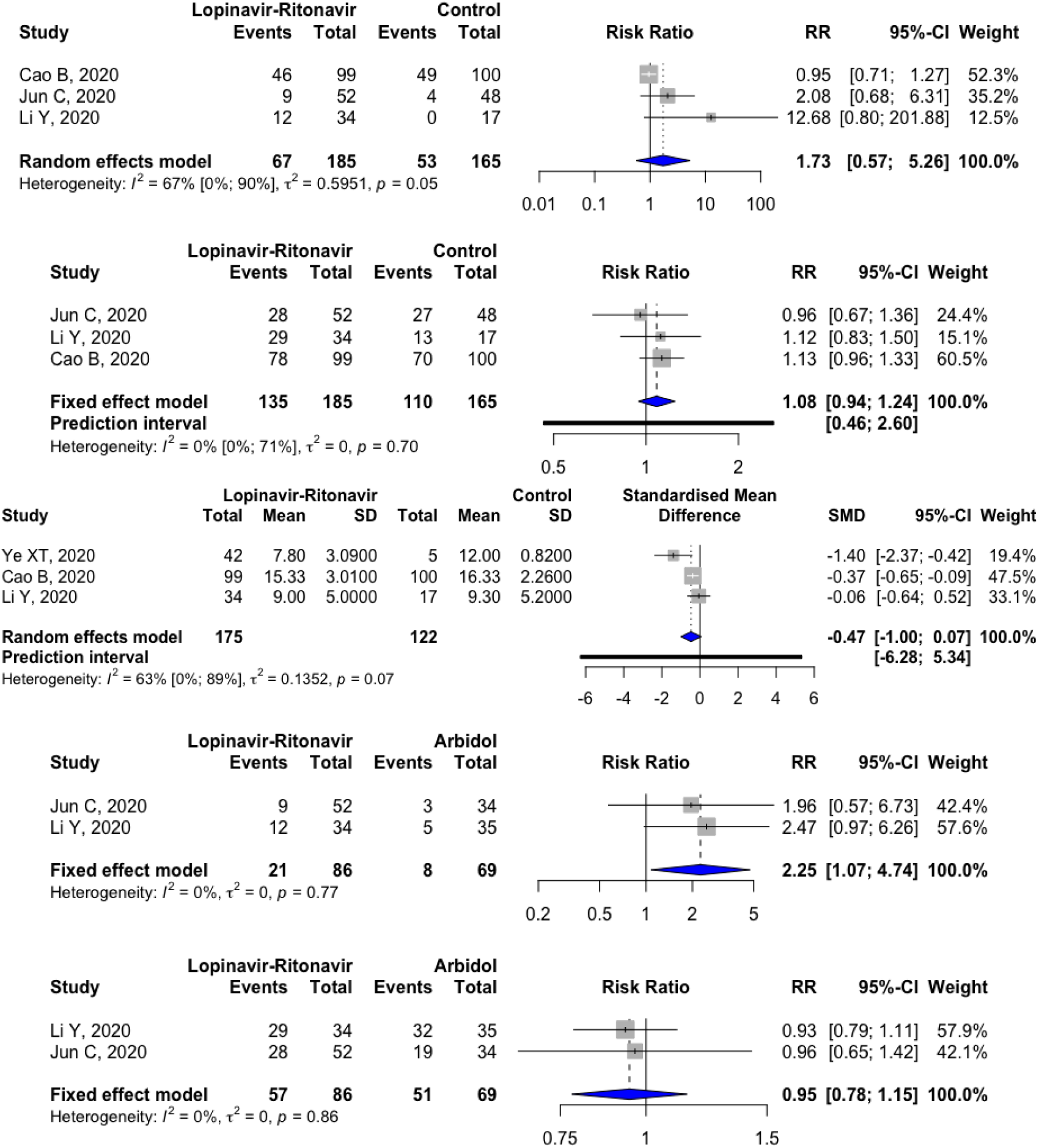
Meta-analysis of lopinavir-ritonavir vs. control groups to assess (a) total adverse events, (b) overall clinical improvement, (c) time to clinical improvement. (d, e): Meta-analysis of lopinavir-ritonavir vs. arbidol groups to assess (d) total adverse events, (e) overall clinical improvement.

#### Lopinavir-Ritonavir Versus Control groups

Four studies consisting of 397 nCOV-2019 cases were included in the meta-analysis and were divided into two groups: 227 subjects to Lopinavir-Ritonavir group and 170 subjects control group. There was no significant association between the two groups in terms of total adverse events (RR 1.73; 95%CI 0.57 to 5.26) and overall clinical improvement (RR 1.08; 95%CI 0.94 to 1.24). Labbé plot observed a trend of having more adverse events towards the Lopinavir-Ritonavir group (supplementary file S2). A borderline association was observed depicting a trend in terms of a shorter mean time (in days) to clinical improvement in the Lopinavir-Ritonavir group compared to the control group (SMD -0.47; 95%CI -1.00 to 0.07) (Fig 3 (a-c)) Due to less number of available studies, a meta-analysis could not be performed for assessing the all-cause mortality between the two groups.

#### Lopinavir-Ritonavir Versus Arbidol groups

The benefit/harm of Lopinavir-Ritonavir treatment over Arbidol treatment was assessed in two studies consisting of 155 nCOV-2019 cases; 86 in the lopinavir-ritonavir treatment group and 69 in Arbidol treatment group. Lopinavir-Ritonavir treatment group was significantly associated with higher total adverse events as compared to the Arbidol treatment group (RR 2.25; 95%CI 1.07 to 4.74). None of the two treatment groups were found to be associated with an increase in the overall clinical improvement of nCOV-2019 patients (RR 0.95; 95%CI 0.78 to 1.15) (Fig 3 (d-e)). The findings were concurrent when analysed using the Labbé plots (supplementary file S2). A meta-analysis could not be performed for all-cause mortality and time to clinical improvement because of less number of studies.

#### Arbidol Versus Control groups

Two studies consisting of 134 nCOV-2019 cases were included in the meta-analysis and were divided into two groups: 69 subjects to Arbidol group and 65 subjects to control group. When compared to the control group, treatment with Arbidol was not found to be significantly associated with either an increase/decrease in the total adverse events (RR 1.80; 95%CI 0.52 to 6.19) or an increase/decrease in the overall clinical improvement (RR 1.08; 95%CI 0.85 to 1.38) (Fig 4 (a, b)). The findings were concurrent when analysed using the Labbé plots (supplementary file S2). A meta-analysis could not be performed for the remaining outcome measures of all-cause mortality and time to clinical improvement due to fewer number of studies.

**Figure 4(a, b):**
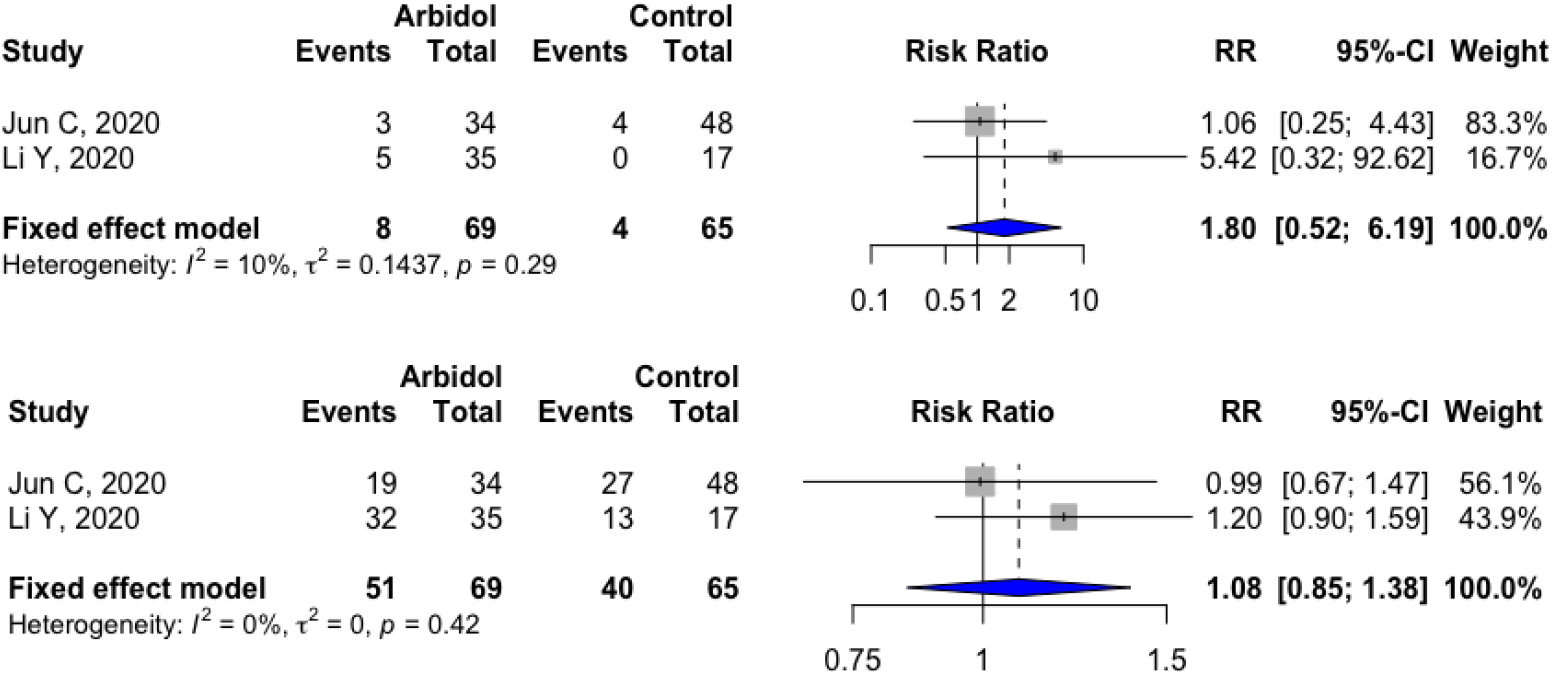
Meta-analysis of arbidol vs. control groups to assess (a) total adverse events, (b) overall clinical improvement.

#### Remdesivir Versus Placebo group

The effect of Remdesivir treatment over placebo was assessed in two RCTs consisting of 1295 nCOV-2019 patients; 696 in Remdesivir group while 599 in placebo group. Compared to placebo group, Remdesivir was not associated with either all-cause mortality (RR 0.74; 95%CI 0.40 to 1.37), total adverse events (RR 0.91; 95%CI 0.79 to 1.05) or time to clinical improvement (SMD -0.78; 95%CI -2.05 to 0.50). However, a significant association was observed with better overall clinical improvement (RR 1.17; 95%CI 1.07 to 1.29) in Remdesivir group compared to placebo group (Figure 5 (a-d)). Similar findings were observed in Labbé plot analysis as well (supplementary file S2).

**Figure 5(a-d):**
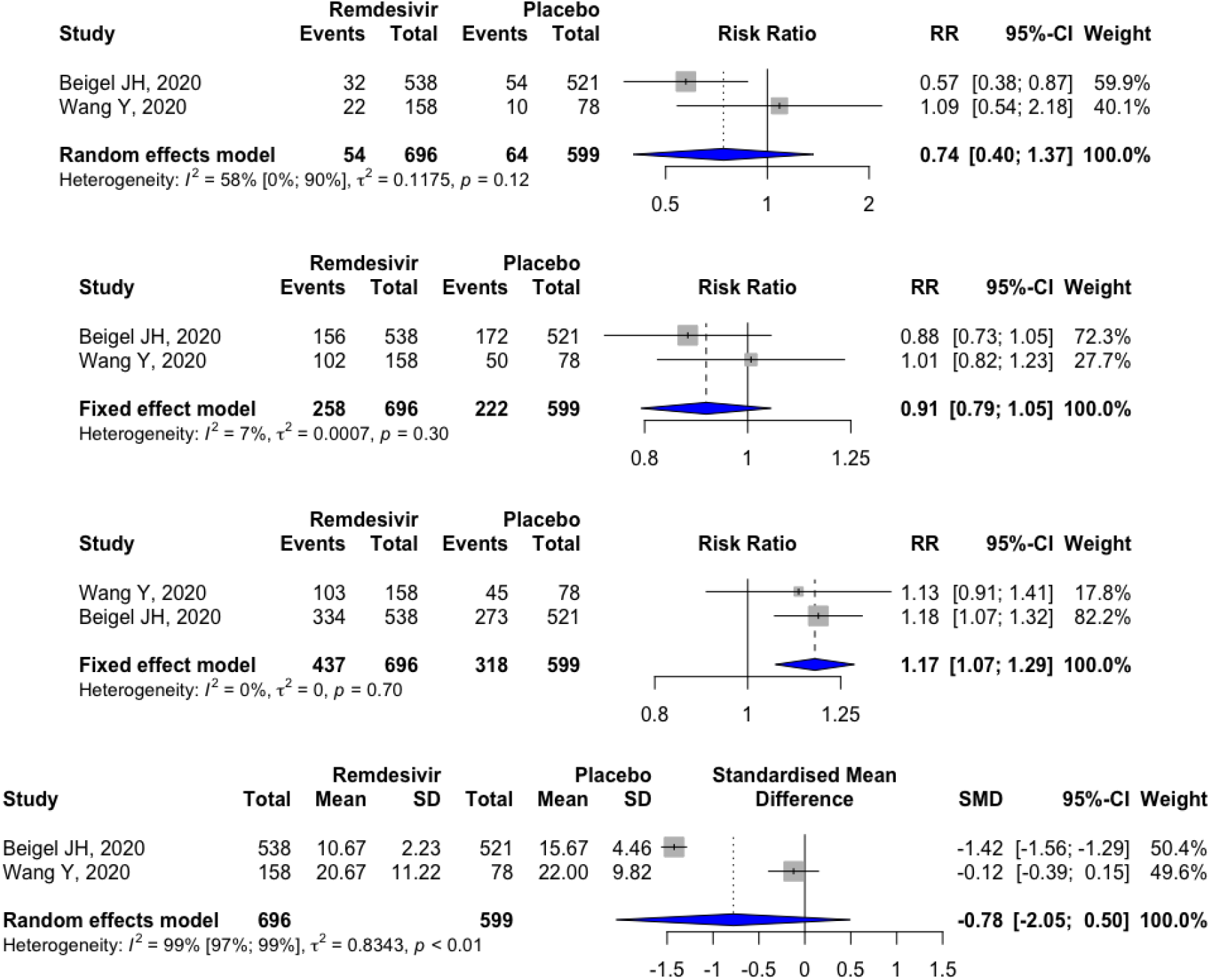
Meta-analysis of Remdesivir vs. Control groups to assess (a) allcause mortality, (b) total adverse events, (c) overall clinical improvement, (d) time to clinical improvement.

#### Corticosteroids Versus Control groups

Four studies consisting of 211 nCOV-2019 cases were included in the meta-analysis and were divided into two groups: 119 subjects to Corticosteroid group and 92 subjects to control group. Administration of Corticosteroid treatment was found to have a borderline association depicting a trend towards an increase in the risk of all-cause mortality as compared to the control group (RR 2.24; 95%CI 0.96 to 5.25). Labbé plots also observed a similar trend of higher all-cause mortality towards the corticosteroid group (supplementary file S2). The Corticosteroid treatment was found to have no significant association with an increase/decrease in the average time to clinical improvement (SMD 0.16; 95%CI -0.26 to 0.58) compared to the control group (Fig 6 (a, b)) Since enough studies could not be pooled, no meta-analysis was performed to assess the effect of Corticosteroid treatment on the total adverse events and overall clinical improvement.

**Figure 6(a, b):**
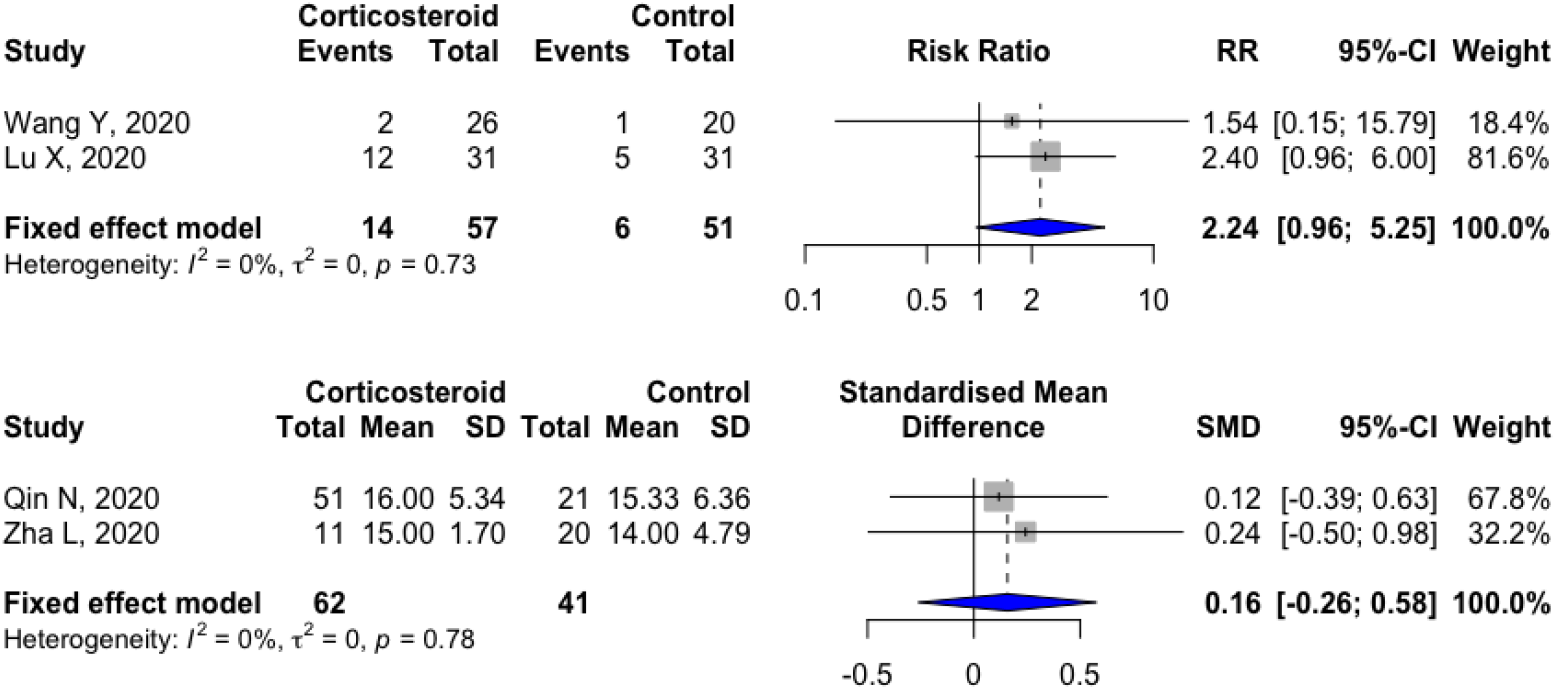
Meta-analysis of Corticosteroid vs. Control groups to assess (a) allcause mortality, (b) time to clinical improvement.

#### Combination therapy

The combination of Hydroxychloroquine and Azithromycin was tested in three studies including 1253 nCOV-2019 cases wherein, 854 cases were allocated to the Hydroxychloroquine + Azithromycin treatment group and the rest 399 cases to the control group. The combination of Hydroxychloroquine + Azithromycin was significantly associated with a higher risk of all-cause mortality compared to the control group (RR 2.01; 95%CI 1.47 to 2.73) while no association was observed between the two in terms of overall clinical improvement (RR 0.95; 95%CI 0.74 to 1.21) (Fig 7 (a, b)). However, we did observe a slight trend in a lesser overall clinical improvement towards the Hydroxychloroquine + Azithromycin treatment group using the Labbé plot (supplementary file S2). The total adverse events and time to clinical improvement outcomes could not be assessed due to limited number of studies.

**Figure 7(a-b):**
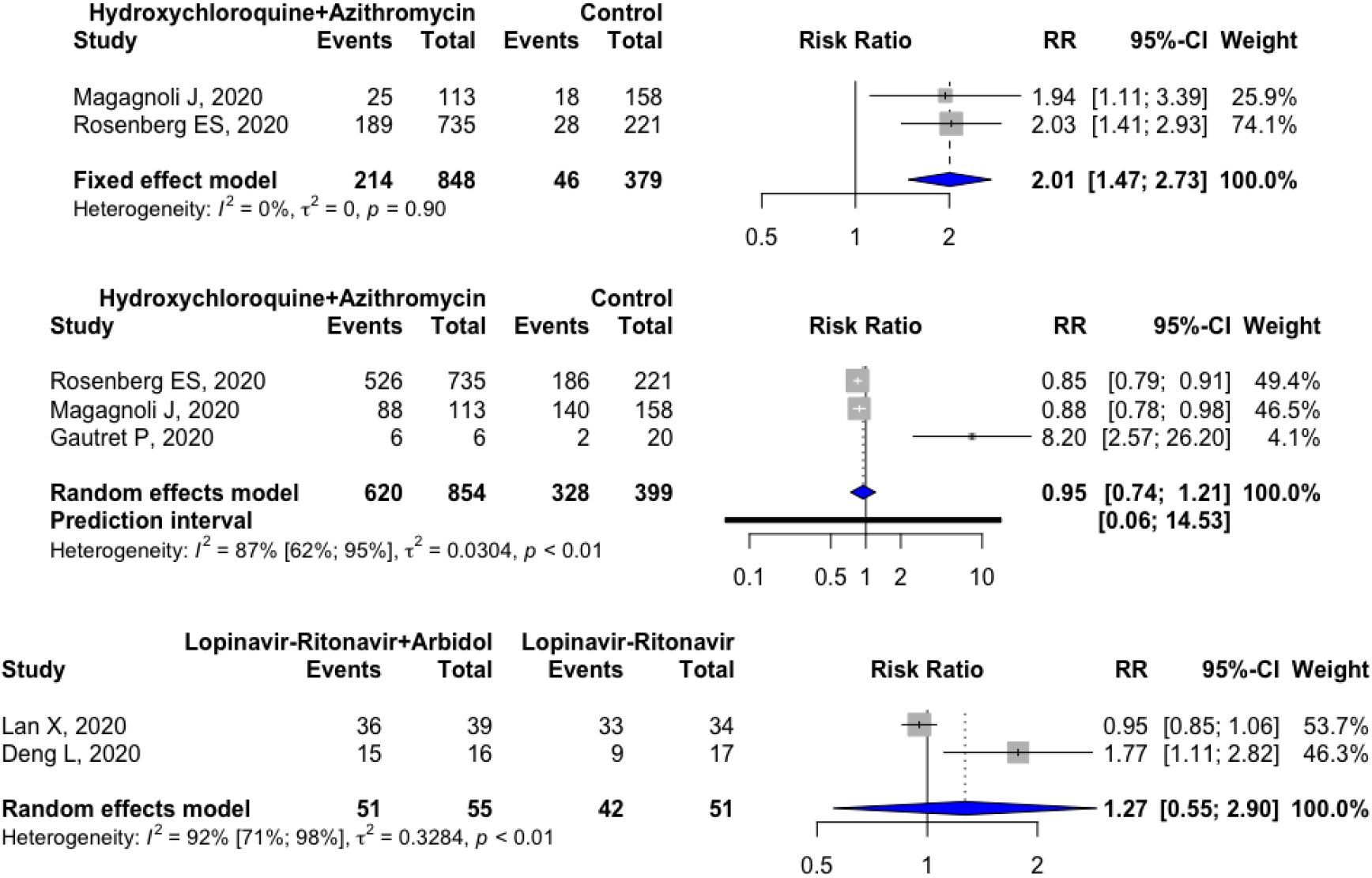
Meta-analysis of combination therapy. Meta-analysis of Hydroxychloroquine + Azithromycin vs. Control groups to assess (a) all-cause mortality, (b) overall clinical improvement. (c):Meta-analysis of combination therapy. Meta-analysis of Lopinavir-Ritonavir + Arbidol vs. Lopinavir-Ritonavir groups to assess (d) overall clinical improvement.

Two studies including 106 nCOV-2019 cases investigated another combination of Lopinavir-Ritonavir + Arbidol in 55 cases compared to Lopinavir-Ritonavir alone in 51 cases. Due to limited number of available studies, the data could only be pooled to assess the overall clinical improvement outcome. The addition of Arbidol to Lopinavir-Ritonavir treatment was not found to be significantly associated with an increase/decrease in the overall clinical improvement (RR 1.27; 95%CI 0.55 to 2.90) compared to Lopinavir-Ritonavir treatment alone (Fig 7 (c))

### Publication bias

The publication bias was assessed using funnel plot analysis for all those treatment modalities wherein data from more than two studies could be pooled together. The shape of the funnel plots did not show any evidence of significant publication bias except for one instance wherein Lopinavir-Ritonavir treatment was compared with control group to assess the total adverse event outcome. This was confirmed by the significant p-value obtained from the Egger’s regression test (p-value: 0.02). The p-value of Egger’s regression test was not significant for the presence of any publication bias for the rest of the funnel plots. Fig 8(b) represents the funnel plot analysis carried out for hydroxychloroquine vs. control group for assessing the overall clinical improvement outcome. The remaining funnel plots have been depicted in the Table S2.1 of supplementary file S2.

**Figure 8(a):**
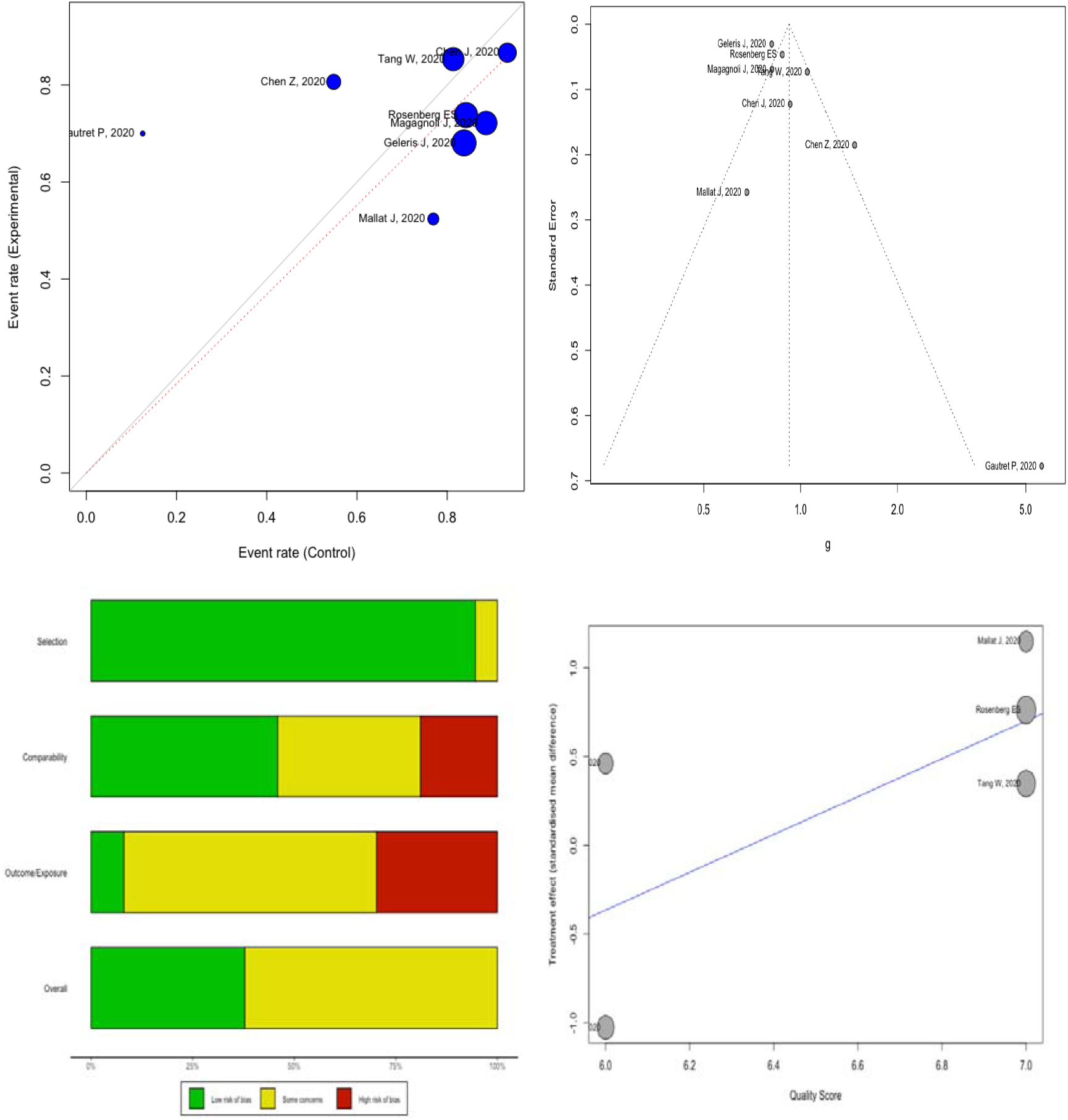
Labbé plot analysis for observing the trend and between-study heterogeneity in meta-analysis between Hydroxychloroquine and Control group for assessing the overall clinical improvement outcome. Figure 8(b): Funnel plot for publication bias analysis of Hydroxychloroquine Vs. Control group assessing the overall clinical improvement (Egger’s P-value: 0.07). Figure 8(c): Risk of bias assessment of the included studies in the systematic review using the Newcastle Ottawa Scale. Figure 8(d): Meta-regression analysis for assessing the time to clinical improvement outcome between Hydroxychloroquine and Control group.

### Quality assessment

The risk of bias in the studies included in our systematic review was assessed using the Newcastle-Ottawa Scale for quality measurement. The total score was divided into three categories: (1) 1-3 (High risk of bias); (2) 4-6 (Some concerns); 7-9 (Low risk of bias). Twelve studies [32.43%] included in our review had an overall low risk of bias while the rest 25 studies [67.57%] had some concerns related to the risk of bias. No study included in the systematic review had an overall high risk of bias (Fig 8 (c)). The individual items for the quality scale are depicted supplementary file S1.

### Meta-regression analysis

A meta-regression analysis was conducted to determine if the quality score of each study was associated with the overall effect size difference. A meta-regression analysis was performed only for those treatments where data from more than two studies was pooled. The “quality score” variable as a predictor, was found to be significantly associated with the overall effect size difference while assessing the time to clinical improvement outcome between Hydroxychloroquine and control groups (P-value: 0.02) (Fig 8 (d)) and similar outcome between Lopinavir-Ritonavir and control groups (P-value: 0.03). Quality score did not have any association with the overall effect size difference for assessing the remaining outcomes between other treatment groups.

### Influence diagnostics and sensitivity analysis

Influence diagnostics tools and sensitivity analysis were used to further explain the heterogeneity observed in our results and to identify the outlier studies which could be significantly affecting the overall pooled effect estimates. The influence diagnostics and sensitivity analysis were performed for treatments in which data from more than two studies was pooled. The influence diagnostic tools generated two plots including 1) Baujat plots; 2) Influence analysis plots; and two plots for sensitivity analysis including 3) leave-one-out analysis ordered by heterogeneity and 4) leave-one-out analysis ordered by effect size.

The Baujat and influence analysis plots identified two potential outliers namely, Magagnoli J, 2020 and Yu B, 2020 while assessing the all-cause mortality outcome in Hydroxychloroquine vs. control group analysis. After conducting the sensitivity analysis by omitting a single study in each turn (ordered by both effect size and I^2^), the overall effect size changed significantly by omitting the Yu B, 2020 study. Hydroxychloroquine was found to be significantly associated with the risk of having more all-cause mortality compared to control group (RR 1.6; 95%CI 1.26 to 2.03). While assessing the overall clinical improvement in the same two groups, the Baujat plot found Geleris J, 2020; Tang W, 2020; Chen Z, 2020 and Gautret P, 2020 as potential outliers. After omitting each study in the sensitivity analysis, we observed that Hydroxychloroquine was significantly associated with less overall clinical improvement than the control group (RR 0.88; 95%CI 0.79 to 0.98) when Chen Z, 2020 study was removed from the pooled analysis (Fig 9). Similarly, after omitting the same Chen Z, 2020 outlier study in the sensitivity analysis for assessing the time to clinical improvement between the two groups, we observed that Hydroxychloroquine was significantly associated with a longer mean time to clinical improvement than the control group (SMD 0.64; 95%CI 0.33 to 0.94).

**Figure 9:**
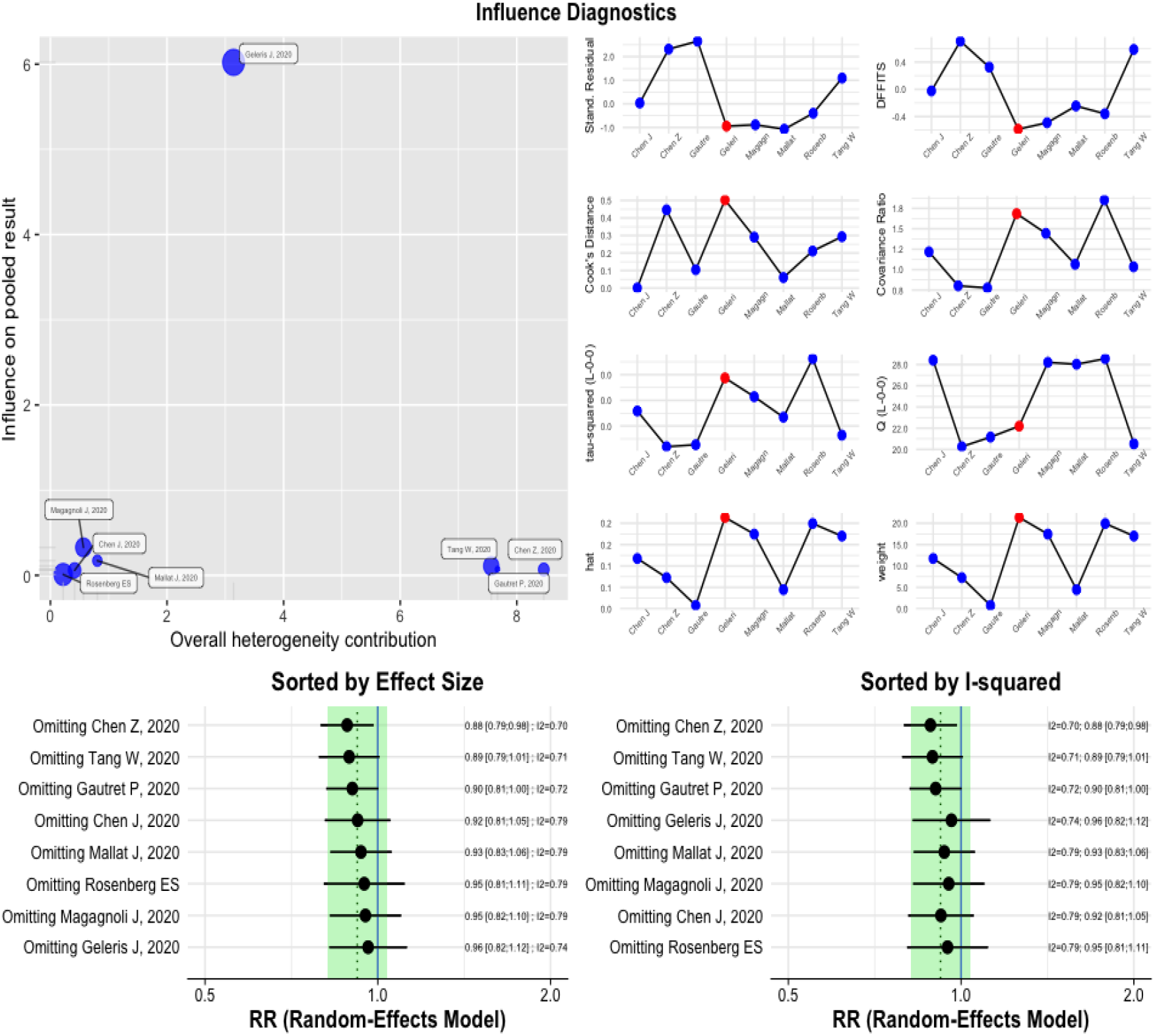
Influence diagnostic tools and sensitivity analysis plots for identifying the potential outlier studies and assessing the heterogeneity in the pooled effect size for assessing the overall clinical improvement outcome between hydroxychloroquine treatment and control group.

The influence diagnostic tools identified Ye XT, 2020 study as a potential outlier while assessing the time to clinical improvement outcome between Lopinavir-Ritonavir and control groups; and after omitting this study in the sensitivity analysis we observed that Lopinavir-Ritonavir was significantly associated with a shorter mean time to clinical improvement than the control group (SMD -0.32; 95%CI -0.57 to -0.06).

Gautret P, 2020 was identified as a significant outlier using the Baujat plot when assessing the overall clinical improvement between Hydroxychloroquine + Azithromycin and control groups. When this study was omitted in the sensitivity analysis, then the combination of Hydroxychloroquine + Azithromycin treatment was found to be significantly associated with lesser overall clinical improvement compared to the control group (RR 0.86; 95%CI 0.81 to 0.91).

## Discussion

Our systematic review and meta-analysis summarized the *in-vitro* and clinical evidence generated so far regarding the effect of various treatment modalities administered to nCOV-2019 pneumonia patients. *In vitro* studies observed significant inhibitory effects of Remdesivir, Hydroxychloroquine and Nelfinavir on nCOV-2019. Hydroxychloroquine was found to have better efficacy and less cytotoxicity than Chloroquine in inhibiting nCOV-2019. However, the clinical translation of promising *in vitro* results in some of these drugs has not been successful. In 37 clinical studies consisting of 8662 nCOV-2019 patients, we assessed the potential of several treatments against their comparators in terms of harm which included all-cause mortality and total adverse events and in terms of benefit which included overall clinical improvement and time to clinical improvement. While assessing the benefits of administered treatments, our meta-analysis observed that Lopinavir-Ritonavir treatment had a borderline association with shorter mean time to clinical improvement compared to the control group while Remdesivir treatment had significant association with better overall clinical improvement compared to placebo group. However, the present evidence stems from only a couple of trials conducted on these drugs and the clinical usefulness of these results will only be determined once further large RCTs are published on the same. In terms of harm, our meta-analysis suggests that Hydroxychloroquine treatment compared to control group and Lopinavir-Ritonavir treatment compared to Arbidol treatment were significantly associated with more total adverse events in nCOV-2019 patients. Hydroxychloroquine combined with Azithromycin was associated with higher all-cause mortality compared to control group. We did not observe any significant association in terms of either benefit or harm for the remaining treatments administered to nCOV-2019 patients when analysed against their respective comparator groups. Although our systematic review observed several treatments associated with benefit including Favipiravir, Chloroquine, Zinc sulphate + Hydroxychloroquine + Azithromycin; and certain treatments associated with harm including corticosteroids and combination therapy of Lopinavir-Ritonavir + Ribavirin + Interferon beta B1 in nCOV-2019 patients (supplementary file S2), the results were reported only from single studies and thus lacked sufficient statistical power to draw any profound conclusions. Around 67.57% of the studies included in our review had moderate/some concerns related to the risk of bias.

When we conducted the influence and sensitivity analysis, we observed that Hydroxychloroquine was associated with a higher all-cause mortality, less overall clinical improvement and a longer mean time to clinical improvement in nCOV-2019 patients when compared to the control group. Furthermore, Hydroxychloroquine combined with Azithromycin was associated with a lesser overall clinical improvement compared to control group after conducting the sensitivity analysis. The borderline association of Lopinavir-Ritonavir treatment having a shorter mean time to clinical improvement compared to control group was confirmed to be statistically significant after the sensitivity analysis. Our findings are in concordance with a review published in April 2020 by Sanders JM et al. which reviewed the initial pharmacological treatments available for nCOV-2019 and concluded that no available therapy was found to be effective for treating this infection (95).

Initial evidence from *in vitro* and observational studies suggested that Hydroxychloroquine has comparatively faster viral clearance and results in better clinical improvement of nCOV-2019 patients in contrast to control groups (5,64). Further, the combination of Hydroxychloroquine and Azithromycin resulted in 100% clinical improvement in a small open label non-RCT published by Gautret et al. 2020 (6). However, when early results from few RCTs were reported, Hydroxychloroquine no longer had any benefit over standard care and instead was associated with more adverse events and higher mortality rate (10,61). We also conducted a subgroup analysis based on study design which further strengthened this notion. Hydroxychloroquine compared to control group was found to be associated with less overall clinical improvement in non-RCTs/cohort study subgroup while no association was observed in the RCT subgroup (Fig S2.2 in supplementary file S2). A recently published multinational registry analysis on 96,032 nCOV-2019 patients observed that Hydroxychloroquine and Chloroquine were associated with an increased risk of in-hospital mortality (96). Two recent meta-analyses conducted by Ren L et al. 2020 and Wang J et al. 2020 found that patients taking Chloroquine or Hydroxychloroquine had more adverse events compared to patients assigned to placebo group (97,98). Another meta-analysis published last month by Sarma et al. 2020 found no association of Hydroxychloroquine with virological cure, death or clinical worsening and safety in nCOV-2019 patients (99). Similar findings on Hydroxychloroquine with or without azithromycin were observed from another meta-analysis of five trials which although did observe a trend but the results were not found to be statistically significant in terms of negative conversion of nCOV-2019 (Odds Ratio (OR) 1.95; 95%CI 0.19 to 19.73) and reduction in progression rate (OR 0.89 95%CI 0.58 to 1.37) (100). Our meta-analysis along with the subgroup and sensitivity analyses further corroborates these findings.

The effectiveness and safety of corticosteroid treatment in nCOV-2019, SARS and MERS have been investigated in several meta-analyses. Use of corticosteroid treatment was found to be associated with higher mortality (RR 2.11; 95%CI 1.13 to 3.94) in nCOV-2019 and SARS patients in a meta-analysis of 15 studies conducted by Yang Z et al. 2020 (101). While three meta-analyses found that corticosteroid use did not worsen/improve mortality in patients with nCOV-2019, SARS-Cov and MERS-Cov (102-104). Further, the meta-analysis by Li H et al. 2020 also observed a delayed time to virus clearance in the corticosteroid group compared to controls (MD 3.78; 95%CI 1.16 to 6.41) (103). The findings of our meta-analysis are also in line with the previously published meta-analyses on corticosteroids. We did not observe any significant association between corticosteroid treatment and all-cause mortality and time to clinical improvement.

Use of Convalescent Plasma has been shown to be extremely promising in some recently published case-series (105,106). However, we could not assess the benefit/harm of plasma therapy for nCOV-2019 patients in our systematic review and meta-analysis due to the scarcity of available literature and specific inclusion criteria of our review. A recently published systematic review of five studies by Rajendran K et al. 2020 concluded that plasma therapy in nCOV-2019 patients was safe, clinically effective and was associated with a reduced mortality (107). Results from ongoing clinical trials on plasma therapy are awaited and will give us a better insight into the effectiveness of Convalescent Plasma in treating nCOV-2019 patients.

### Limitations

Although we made sure that our systematic review and meta-analysis was conducted very comprehensively, certain inherent and obvious limitations cannot be ignored. Firstly, due to the limited number of studies, our meta-analysis pooled the data from RCTs and non-RCTs/cohort/case-control studies together which is generally not advisable. However, we did conduct a subgroup analysis based on study design wherever possible to separate the RCTs from non-RCTs/cohort/case-control studies. Secondly, all outcome measures could not be assessed for all the potential treatments due to scarcity of literature. Thirdly, a significant amount of studies included in the review were in their preprint versions and yet to be peer reviewed by experts. Lastly, since several clinical trials on nCQV-2019 treatments are currently ongoing, the results of our meta-analysis might change significantly owing to the findings published in near future.

Nonetheless, our meta-analysis presents preliminary evidence of benefit/harm of the possible treatments being administered to nCOV-2019 patients and these preliminary results could be used for conducting and planning large clinical trials and prospective multicentric cohort studies.

## Conclusion

The result of this systematic review and meta-analysis suggest that Hydroxychloroquine, Remdesivir and Nelfinavir have shown promising results in the *in vitro* studies. However, based on the current clinical evidence, our meta-analysis did not observe significant beneficial effect of any treatment on nCOV-2019 patients apart from a significant association in better overall clinical improvement of Remdesivir compared to placebo and a borderline association in time to clinical improvement of Lopinavir-Ritonavir treatment compared to control group. Hydroxychloroquine with or without azithromycin might be associated with higher all-cause mortality, more total adverse events, less overall clinical improvement and a higher mean time to clinical improvement. Results from further large clinical trials are warranted.

## Data Availability

The above manuscript is a systematic review and meta-analysis and the data individual study data has been collected from the articles. The raw data for meta-analysis may be available upon request for the critical appraisal of the manuscript.

## Acknowledgement

None

## References

1. Zhu N, Zhang D, Wang W, Li X, Yang B, Song J, et al. A Novel Coronavirus from Patients with Pneumonia in China, 2019. N Engl J Med. 2020 20;382(8):727–33.

2. Dong E, Du H, Gardner L. An interactive web-based dashboard to track COVID-19 in real time. Lancet Infect Dis [Internet]. 2020 Feb 19 [cited 2020 Mar 28];0(0). Available from: https://www.thelancet.com/journals/laninf/article/PIIS1473-3099(20)30120-1/abstract

3. de Wit E, van Doremalen N, Falzarano D, Munster VJ. SARS and MERS: recent insights into emerging coronaviruses. Nat Rev Microbiol. 2016;14(8):523–34.

4. Wang M, Cao R, Zhang L, Yang X, Liu J, Xu M, et al. Remdesivir and chloroquine effectively inhibit the recently emerged novel coronavirus (2019-nCoV) in vitro. Cell Res. 2020;30(3):269–71.

5. Yao X, Ye F, Zhang M, Cui C, Huang B, Niu P, et al. In Vitro Antiviral Activity and Projection of Optimized Dosing Design of Hydroxychloroquine for the Treatment of Severe Acute Respiratory Syndrome Coronavirus 2 (SARS-CoV-2). Clin Infect Dis Off Publ Infect Dis Soc Am. 2020 Mar 9;

6. Gautret P, Lagier J-C, Parola P, Hoang VT, Meddeb L, Mailhe M, et al. Hydroxychloroquine and azithromycin as a treatment of COVID-19: results of an open-label non-randomized clinical trial. Int J Antimicrob Agents. 2020 Mar 20;105949.

7. NIH Clinical Trial Shows Remdesivir Accelerates Recovery from Advanced COVID-19 | NIH: National Institute of Allergy and Infectious Diseases [Internet], [cited 2020 May 21], Available from: http://www.niaid.nih.gov/news-events/nih-clinical-trial-shows-remdesivir-accelerates-recovery-advanced-covid-19

8. Vincent AL. EUA Hydroxychloroquine sulfate Health Care Provider Fact Sheet 04272020.:7.

9. Commissioner O of the. Coronavirus (COVID-19) Update: FDA Issues Emergency Use Authorization for Potential COVID-19 Treatment [Internet]. FDA. FDA; 2020 [cited 2020 May 21], Available from: https://www.fda.gov/news-events/press-announcements/coronavirus-covid-19-update-fda-issues-emergency-use-authorization-potential-covid-19-treatment

10. Tang W, Cao Z, Han M, Wang Z, Chen J, Sun W, et al. Hydroxychloroquine in patients with mainly mild to moderate coronavirus disease 2019: open label, randomised controlled trial. BMJ. 2020 14;369:m1849.

11. Wang Y, Zhang D, Du G, Du R, Zhao J, Jin Y, et al. Remdesivir in adults with severe COVID-19: a randomised, double-blind, placebo-controlled, multicentre trial. Lancet Lond Engl. 2020 May 16;395(10236):1569–78.

12. Research C for DE and. FDA cautions against use of hydroxychloroquine or chloroquine for COVID-19 outside of the hospital setting or a clinical trial due to risk of heart rhythm problems. FDA [Internet]. 2020 Apr 30 [cited 2020 May 21]; Available from: https://www.fda.gov/drugs/drug-safety-and-availability/fda-cautions-against-use-hydroxychloroquine-or-chloroquine-covid-19-outside-hospital-setting-or

13. Moher D, Shamseer L, Clarke M, Ghersi D, Liberati A, Petticrew M, et al. Preferred reporting items for systematic review and meta-analysis protocols (PRISMA-P) 2015 statement. Syst Rev. 2015 Jan 1;4:1.

14. Ottawa Hospital Research Institute [Internet], [cited 2020 May 21]. Available from: http://www.ohri.ca/programs/clinical_epidemiology/oxford.asp

15. Egger M, Davey Smith G, Schneider M, Minder C. Bias in meta-analysis detected by a simple, graphical test. BMJ. 1997 Sep 13;315(7109):629–34.

16. Wan X, Wang W, Liu J, Tong T. Estimating the sample mean and standard deviation from the sample size, median, range and/or interquartile range. BMC Med Res Methodol. 2014 Dec 19;14:135.

17. Wang X, Cao R, Zhang H, Liu J, Xu M, Hu H, et al. The anti-influenza virus drug, arbidol is an efficient inhibitor of SARS-CoV-2 in vitro. Cell Discov. 2020 May 2;6(1):1–5.

18. Su H, Yao S, Zhao W, Li M, Liu J, Shang W, et al. Discovery of baicalin and baicalein as novel, natural product inhibitors of SARS-CoV-2 3CL protease in vitro. bioRxiv. 2020 Apr 14;2020.04.13.038687.

19. Kim H-Y, Shin H-S, Park H, Kim Y-C, Yun YG, Park S, et al. In vitro inhibition of coronavirus replications by the traditionally used medicinal herbal extracts, Cimicifuga rhizoma, Meliae cortex, Coptidis rhizoma, and Phellodendron cortex. J Clin Virol. 2008 Feb 1;41(2):122–8.

20. Wu Y, Li C, Xia S, Tian X, Wang Z, Kong Y, et al. Fully human single-domain antibodies against SARS-CoV-2. bioRxiv. 2020 Mar 31;2020.03.30.015990.

21. Yao X, Ye F, Zhang M, Cui C, Huang B, Niu P, et al. In Vitro Antiviral Activity and Projection of Optimized Dosing Design of Hydroxychloroquine for the Treatment of Severe Acute Respiratory Syndrome Coronavirus 2 (SARS-CoV-2). Clin Infect Dis [Internet], [cited 2020 May 21]; Available from: https://academic.oup.com/cid/advance-article/doi/10.1093/cid/ciaa237/5801998

22. Liu J, Cao R, Xu M, Wang X, Zhang H, Hu H, et al. Hydroxychloroquine, a less toxic derivative of chloroquine, is effective in inhibiting SARS-CoV-2 infection in vitro. Cell Discov. 2020 Mar 18;6(1):1–4.

23. Yang J, Wu M, Liu X, Liu Q, Guo Z, Yao X, et al. Cytotoxicity evaluation of chloroquine and hydroxychloroquine in multiple cell lines and tissues by dynamic imaging system and PBPK model. bioRxiv. 2020 Apr 27;2020.04.22.056762.

24. Xu T, Gao X, Wu Z, Selinger DW, Zhou Z. Indomethacin has a potent antiviral activity against SARS CoV-2 in vitro and canine coronavirus in vivo. bioRxiv. 2020 Apr 5;2020.04.01.017624.

25. Runfeng L, Yunlong H, Jicheng H, Weiqi P, Qinhai M, Yongxia S, et al. Lianhuaqingwen exerts anti-viral and anti-inflammatory activity against novel coronavirus (SARS-CoV-2). Pharmacol Res. 2020 Jun 1;156:104761.

26. Zheng F, Zhou Y, Zhou Z, Ye F, Huang B, Huang Y, et al. A Novel Protein Drug, Novaferon, as the Potential Antiviral Drug for COVID-19. medRxiv. 2020 Apr 29;2020.04.24.20077735.

27. Gu C, Wu Y, Guo H, Zhu Y, Xu W, Wang Y, et al. Potent antiviral effect of protoporphyrin IX and verteporfin on SARS-CoV-2 infection. bioRxiv. 2020 May 1;2020.04.30.071290.

28. Lu S, Pan X, Chen D, Xie X, Wu Y, Shang W, et al. Broad-spectrum antivirals of protoporphyrins inhibit the entry of highly pathogenic emerging viruses. bioRxiv. 2020 May 10;2020.05.09.085811.

29. Deng W, Xu Y, Kong Q, Xue J, Yu P, Liu J, et al. Therapeutic efficacy of Pudilan Xiaoyan Oral Liquid (PDL) for COVID-19 in vitro and in vivo. Signal Transduct Target Ther. 2020 May 8;5(l):l-3.

30. Wang M, Cao R, Zhang L, Yang X, Liu J, Xu M, et al. Remdesivir and chloroquine effectively inhibit the recently emerged novel coronavirus (2019-nCoV) in vitro. Cell Res. 2020 Mar;30(3):269–71.

31. Choy K-T, Wong AY-L, Kaewpreedee P, Sia SF, Chen D, Hui KPY, et al. Remdesivir, lopinavir, emetine, and homoharringtonine inhibit SARS-CoV-2 replication in vitro. Antiviral Res. 2020 Jun 1;178:104786.

32. Rothan HA, Stone S, Natekar J, Kumari P, Arora K, Kumar M. The FDA-approved gold drug Auranofin inhibits novel coronavirus (SARS-COV-2) replication and attenuates inflammation in human cells. bioRxiv. 2020 Apr 15;2020.04.14.041228.

33. Sheahan TP, Sims AC, Zhou S, Graham RL, Pruijssers AJ, Agostini ML, et al. An orally bioavailable broad-spectrum antiviral inhibits SARS-CoV-2 in human airway epithelial cell cultures and multiple coronaviruses in mice. Sci Transl Med [Internet]. 2020 Apr 29 [cited 2020 May 21];12(541). Available from: https://stm.sciencemag.org/content/12/541/eabb5883

34. Ma C, Sacco MD, Hurst B, Townsend JA, Hu Y, Szeto T, et al. Boceprevir, GC-376, and calpain inhibitors II, XII inhibit SARS-CoV-2 viral replication by targeting the viral main protease. bioRxiv. 2020 May 8;2020.04.20.051581.

35. Mantlo E, Bukreyeva N, Maruyama J, Paessler S, Huang C. Antiviral activities of type I interferons to SARS-CoV-2 infection. Antiviral Res. 2020 Jul 1;179:104811.

36. Pruijssers AJ, George AS, Schäfer A, Leist SR, Gralinksi LE, Dinnon KH, et al. Remdesivir potently inhibits SARS-CoV-2 in human lung cells and chimeric SARS-CoV expressing the SARS-CoV-2 RNA polymerase in mice. bioRxiv. 2020 Apr 27;2020.04.27.064279.

37. Liu S, Lien CZ, Selvaraj P, Wang TT. Evaluation of 19 antiviral drugs against SARS-CoV-2 Infection. bioRxiv. 2020 May 7;2020.04.29.067983.

38. Touret F, Gilles M, Barral K, Nougairède A, Decroly E, Lamballerie X de, et al. In vitro screening of a FDA approved chemical library reveals potential inhibitors of SARS-CoV-2 replication. bioRxiv. 2020 Apr 5;2020.04.03.023846.

39. Plaze M, Attali D, Prot M, Petit A-C, Blatzer M, Vinckier F, et al. Inhibition of the replication of SARS-CoV-2 in human cells by the FDA-approved drug chlorpromazine. bioRxiv. 2020 May 6;2020.05.05.079608.

40. Terrier O, Dilly S, Pizzorno A, Henri J, Berenbaum F, Lina B, et al. Broad-spectrum antiviral activity of naproxen: from Influenza A to SARS-CoV-2 Coronavirus. bioRxiv. 2020 May 1;2020.04.30.069922.

41. Pizzorno A, Padey B, Julien T, Trouillet-Assant S, Traversier A, Errazuriz-Cerda E, et al. Characterization and treatment of SARS-CoV-2 in nasal and bronchial human airway epithelia. bioRxiv. 2020 Apr 2;2020.03.31.017889.

42. Shannon A, Selisko B, Le NTT, Huchting J, Touret F, Piorkowski G, et al. Favipiravir strikes the SARS-CoV-2 at its Achilles heel, the RNA polymerase. bioRxiv. 2020 May 15;2020.05.15.098731.

43. Matsuyama S, Kawase M, Nao N, Shirato K, Ujike M, Kamitani W, et al. The inhaled corticosteroid ciclesonide blocks coronavirus RNA replication by targeting viral NSP15. bioRxiv. 2020 Mar 12;2020.03.11.987016.

44. Yamamoto M, Kiso M, Sakai-Tagawa Y, Iwatsuki-Horimoto K, Imai M, Takeda M, et al. The anticoagulant nafamostat potently inhibits SARS-CoV-2 infection in vitro: an existing drug with multiple possible therapeutic effects. bioRxiv. 2020 Apr 23;2020.04.22.054981.

45. Yamamoto N, Matsuyama S, Hoshino T, Yamamoto N. Nelfinavir inhibits replication of severe acute respiratory syndrome coronavirus 2 in vitro. bioRxiv. 2020 Apr 8;2020.04.06.026476.

46. Ohashi H, Watashi K, Saso W, Shionoya K, Iwanami S, Hirokawa T, et al. Multidrug treatment with nelfinavir and cepharanthine against COVID-19. bioRxiv. 2020 Apr 15;2020.04.14.039925.

47. Meyer SD, Bojkova D, Cinati J, Damme EV, Buyck C, Loock MV, et al. Lack of Antiviral Activity of Darunavir against SARS-CoV-2. medRxiv. 2020 Apr 8;2020.04.03.20052548.

48. Gassen NC, Papies J, Bajaj T, Dethloff F, Emanuel J, Weckmarm K, et al. Analysis of SARS-CoV-2-controlled autophagy reveals spermidine, MK-2206, and niclosamide as putative antiviral therapeutics. bioRxiv. 2020 Apr 15;2020.04.15.997254.

49. Wang C, Li W, Drabek D, Okba NMA, Haperen R van, Osterhaus ADME, et al. A human monoclonal antibody blocking SARS-CoV-2 infection. Nat Commun. 2020 May 4;11(1):1–6.

50. Silva CSB da, Thaler M, Tas A, Ogando NS, Bredenbeek PJ, Ninaber DK, et al. Suramin inhibits SARS-CoV-2 infection in cell culture by interfering with early steps of the replication cycle. bioRxiv. 2020 May 7;2020.05.06.081968.

51. Jeon S, Ko M, Lee J, Choi I, Byun SY, Park S, et al. Identification of antiviral drug candidates against SARS-CoV-2 from FDA-approved drugs. bioRxiv. 2020 Mar 28;2020.03.20.999730.

52. Ko M, Jeon S, Ryu W-S, Kim S. Comparative analysis of antiviral efficacy of FDA-approved drugs against SARS-CoV-2 in human lung cells: Nafamostat is the most potent antiviral drug candidate. bioRxiv. 2020 May 12;2020.05.12.090035.

53. Caly L, Druce JD, Catton MG, Jans DA, Wagstaff KM. The FDA-approved drug ivermectin inhibits the replication of SARS-CoV-2 in vitro. Antiviral Res. 2020 Jun 1;178:104787.

54. Fintelman-Rodrigues N, Sacramento CQ, Lima CR, Silva FS da, Ferreira AC, Mattos M, et al. Atazanavir inhibits SARS-CoV-2 replication and pro-inflammatory cytokine production. bioRxiv. 2020 Apr 6;2020.04.04.020925.

55. Vuong W, Khan MB, Fischer C, Arutyunova E, Lamer T, Shields J, et al. Feline coronavirus drug inhibits the main protease of SARS-CoV-2 and blocks virus replication. bioRxiv. 2020 May 5;2020.05.03.073080.

56. Vitner EB, Avraham R, Achdout H, Tamir H, Agami A, Cherry L, et al. Antiviral activity of Glucosylceramide synthase inhibitors against SARS-CoV-2 and other RNA virus infections. bioRxiv. 2020 May 19;2020.05.18.103283.

57. Rajasekharan S, Bonotto RM, Kazungu Y, Alves LN, Poggianella M, Orellana PM, et al. Repurposing of Miglustat to inhibit the coronavirus Severe Acquired Respiratory Syndrome SARS-CoV-2. bioRxiv. 2020 May 18;2020.05.18.101691.

58. Ianevski A, Yao R, Fenstad MH, Biza S, Zusinaite E, Lysvand H, et al. Potential antiviral options against SARS-CoV-2 infection. bioRxiv. 2020 May 14;2020.05.12.091165.

59. Holwerda M, V’kovski P, Wider M, Thiel V, Dijkman R. Identification of five antiviral compounds from the Pandemic Response Box targeting SARS-CoV-2. bioRxiv. 2020 May 17;2020.05.17.100404.

60. Weston S, Coleman CM, Haupt R, Logue J, Matthews K, Frieman MB. Broad anti-coronaviral activity of FDA approved drugs against SARS-CoV-2 in vitro and SARS-CoV in vivo. bioRxiv. 2020 Apr 27;2020.03.25.008482.

61. Chen J, Liu D, Liu L, Liu P, Xu Q, Xia L, et al. [A pilot study of hydroxychloroquine in treatment of patients with moderate COVID-19]. Zhejiang Xue Xue Bao Yi Xue Ban J Zhejiang Univ Med Sci. 2020 May 25;49(2):215–9.

62. Chen Z, Hu J, Zhang Z, Jiang S, Han S, Yan D, et al. Efficacy of hydroxychloroquine in patients with COVID-19: results of a randomized clinical trial. medRxiv [Internet]. 2020 Apr 10 [cited 2020 May 20];2020.03.22.20040758. Available from: https://www.medrxiv.org/content/10.1101/2020.03.22.20040758v3

63. Chen C, Zhang Y, Huang J, Yin P, Cheng Z, Wu J, et al. Favipiravir versus Arbidol for COVID-19: A Randomized Clinical Trial. medRxiv [Internet]. 2020 Apr 15 [cited 2020 May 20];2020.03.17.20037432. Available from: https://www.medrxiv.org/content/10.1101/2020.03.17.20037432v4

64. Yu B, Wang DW, Li C. Hydroxychloroquine application is associated with a decreased mortality in critically ill patients with COVID-19. medRxiv [Internet]. 2020 May 1 [cited 2020 May 20];2020.04.27.20073379. Available from: https://www.medrxiv.org/content/10.1101/2020.04.27.20073379vl

65. Huang M, Li M, Xiao F, Liang J, Pang P, Tang T, et al. Preliminary evidence from a multicenter prospective observational study of the safety and efficacy of chloroquine for the treatment of COVID-19. medRxiv [Internet]. 2020 May 4 [cited 2020 May 20];2020.04.26.20081059. Available from: https://www.medrxiv.org/content/10.1101/2020.04.26.20081059vl

66. Huang M, Tang T, Pang P, Li M, Ma R, Lu J, et al. Treating COVID-19 with Chloroquine. J Mol Cell Biol. 2020 18;12(4):322–5.

67. Cao B, Wang Y, Wen D, Liu W, Wang J, Fan G, et al. A Trial of Lopinavir-Ritonavir in Adults Hospitalized with Severe Covid-19. N Engl J Med. 2020 07;382(19):1787–99.

68. Ye X-T, Luo Y-L, Xia S-C, Sun Q-F, Ding J-G, Zhou Y, et al. Clinical efficacy of lopinavir/ritonavir in the treatment of Coronavirus disease 2019. Eur Rev Med Pharmacol Sci. 2020;24(6):3390–6.

69. Jun C, Yun L, Xiuhong X, Ping L, Feng L, Tao L, et al. Efficacies of lopinavir/ritonavir and abidol in the treatment of novel coronavirus pneumonia. Chin J Infect Dis [Internet]. 2020 Feb 21 [cited 2020 May 20];38(00):E008-E008. Available from: http://rs.yiigle.com/yufabiao/1182592.htm

70. Li Y, Xie Z, Lin W, Cai W, Wen C, Guan Y, et al. An exploratory randomized controlled study on the efficacy and safety of lopinavir/ritonavir or arbidol treating adult patients hospitalized with mild/moderate COVID-19 (ELACOI). medRxiv [Internet]. 2020 Apr 15 [cited 2020 May 20];2020.03.19.20038984. Available from: https://www.medrxiv.org/content/10.1101/2020.03.19.20038984v2

71. Cai Q, Yang M, Liu D, Chen J, Shu D, Xia J, et al. Experimental Treatment with Favipiravir for COVID-19: An Open-Label Control Study. Eng Beijing China. 2020 Mar 18;

72. Lou Y, Liu L, Qiu Y. Clinical Outcomes and Plasma Concentrations of Baloxavir Marboxil and Favipiravir in COVID-19 Patients: an Exploratory Randomized, Controlled Trial. medRxiv [Internet]. 2020 May 5 [cited 2020 May 20];2020.04.29.20085761. Available from: https://www.medrxiv.org/content/10.1101/2020.04.29.20085761vl

73. Deng L, Li C, Zeng Q, Liu X, Li X, Zhang H, et al. Arbidol combined with LPV/r versus LPV/r alone against Corona Virus Disease 2019: A retrospective cohort study. J Infect. 2020 Mar 11;

74. Lan X, Shao C, Zeng X, Wu Z, Xu Y. Lopinavir-ritonavir alone or combined with arbidol in the treatment of 73 hospitalized patients with COVID-19: a pilot retrospective study. medRxiv [Internet]. 2020 Apr 29 [cited 2020 May 20];2020.04.25.20079079. Available from: https://www.medrxiv.org/content/10.1101/2020.04.25.20079079vl

75. Wang D, Wang J, Jiang Q, Yang J, Li J, Gao C, et al. No Clear Benefit to the Use of Corticosteroid as Treatment in Adult Patients with Coronavirus Disease 20190: A Retrospective Cohort Study. medRxiv [Internet]. 2020 Apr 24 [cited 2020 May 20];2020.04.21.20066258. Available from: https://www.medrxiv.org/content/10.1101/2020.04.21.20066258vl

76. Wang Y, Jiang W, He Q, Wang C, Wang B, Zhou P, et al. A retrospective cohort study of methylprednisolone therapy in severe patients with COVID-19 pneumonia. Signal Transduct Target Ther. 2020 28;5(1):57.

77. Zha L, Li S, Pan L, Tefsen B, Li Y, French N, et al. Corticosteroid treatment of patients with coronavirus disease 2019 (COVID-19). Med J Aust. 2020;212(9):416–20.

78. Lu X, Chen T, Wang Y, Wang J, Zhang B, Li Y, et al. Adjuvant corticosteroid therapy for critically ill patients with COVID-19. medRxiv [Internet]. 2020 Apr 11 [cited 2020 May 20];2020.04.07.20056390. Available from: https://www.medrxiv.org/content/10.1101/2020.04.07.20056390vl

79. Qin N, Cheng D, Yongtao L, Hong Z, Jun L, Xuan Z, et al. Retrospective study of low-to-moderate dose glucocorticoids on viral clearance in patients with novel coronavirus pneumonia. Chin J Clin Infect Dis [Internet]. 2020 Feb 28 [cited 2020 May 20];13(00):E009-E009. Available from: http://rs.yiigle.com/yufabiao/1182773.htm

80. Shi C, Wang C, Wang H, Yang C, Cai F, Zeng F, et al. The potential of low molecular weight heparin to mitigate cytokine storm in severe COVID-19 patients: a retrospective clinical study. medRxiv [Internet]. 2020 Apr 15 [cited 2020 May 20];2020.03.28.20046144. Available from: https://www.medrxiv.org/content/10.1101/2020.03.28.20046144v3

81. Zhong M, Sun A, Xiao T, Yao G, Sang L, Zheng X, et al. A Randomized, Single-blind, Group sequential, Active-controlled Study to evaluate the clinical efficacy and safety of α-Lipoic acid for critically ill patients with coronavirus disease 2019 (COVID-19). medRxiv [Internet]. 2020 Apr 21 [cited 2020 May 20];2020.04.15.20066266. Available from: https://www.medrxiv.org/content/10.1101/2020.04.15.20066266vl

82. Bian H, Zheng Z-H, Wei D, Zhang Z, Kang W-Z, Hao C-Q, et al. Meplazumab treats COVID-19 pneumonia: an open-labelled, concurrent controlled add-on clinical trial. medRxiv [Internet]. 2020 Mar 24 [cited 2020 May 20];2020.03.21.20040691. Available from: https://www.medrxiv.org/content/10.1101/2020.03.21.20040691vl

83. Liu X, Li Z, Liu S, Chen Z, Zhao Z, Huang Y, et al. Therapeutic effects of dipyridamole on COVID-19 patients with coagulation dysfunction. medRxiv [Internet]. 2020 Feb 29 [cited 2020 May 20];2020.02.27.20027557. Available from: https://www.medrxiv.org/content/10.1101/2020.02.27.20027557vl

84. Geleris J, Sun Y, Platt J, Zucker J, Baldwin M, Hripcsak G, et al. Observational Study of Hydroxychloroquine in Hospitalized Patients with Covid-19. N Engl J Med. 2020 May 7;

85. Magagnoli J, Narendran S, Pereira F, Cummings T, Hardin JW, Sutton SS, et al. Outcomes of hydroxychloroquine usage in United States veterans hospitalized with Covid-19. medRxiv [Internet]. 2020 Apr 23 [cited 2020 May 20];2020.04.16.20065920. Available from: https://www.medrxiv.org/content/10.1101/2020.04.16.20065920v2

86. Rosenberg ES, Dufort EM, Udo T, Wilberschied LA, Kumar J, Tesoriero J, et al. Association of Treatment With Hydroxychloroquine or Azithromycin With In-Hospital Mortality in Patients With COVID-19 in New York State. JAMA. 2020 May 11;

87. Carlucci P, Ahuja T, Petrilli CM, Rajagopalan H, Jones S, Rahimian J. Hydroxychloroquine and azithromycin plus zinc vs hydroxychloroquine and azithromycin alone: outcomes in hospitalized COVID-19 patients. medRxiv [Internet]. 2020 May 8 [cited 2020 May 20];2020.05.02.20080036. Available from: https://www.medrxiv.org/content/10.1101/2020.05.02.20080036vl

88. Fadel R, Morrison AR, Vahia A, Smith ZR, Chaudhry Z, Bhargava P, et al. Early Short Course Corticosteroids in Hospitalized Patients with COVID-19. Clin Infect Dis Off Publ Infect Dis Soc Am. 2020 May 19;

89. Beigel JH, Tomashek KM, Dodd LE, Mehta AK, Zingman BS, Kalil AC, et al. Remdesivir for the Treatment of Covid-19 — Preliminary Report. N Engl J Med [Internet]. 2020 May 22 [cited 2020 May 23];0(0):null. Available from: https://doi.org/10.1056/NEJMoa2007764

90. Mahevas M, Tran V-T, Roumier M, Chabrol A, Paule R, Guillaud C, et al. No evidence of clinical efficacy of hydroxychloroquine in patients hospitalized for COVID-19 infection with oxygen requirement: results of a study using routinely collected data to emulate a target trial. medRxiv [Internet]. 2020 Apr 14 [cited 2020 May 20];2020.04.10.20060699. Available from: https://www.medrxiv.org/content/10.1101/2020.04.10.20060699vl

91. Mallat J, Hamed F, Balkis M, Mohamed MA, Mooty M, Malik A, et al. Hydroxychloroquine is associated with slower viral clearance in clinical COVID-19 patients with mild to moderate disease: A retrospective study. medRxiv [Internet]. 2020 May 2 [cited 2020 May 20];2020.04.27.20082180. Available from: https://www.medrxiv.org/content/10.1101/2020.04.27.20082180vl

92. Borba MGS, Val FFA, Sampaio VS, Alexandre MAA, Melo GC, Brito M, et al. Effect of High vs Low Doses of Chloroquine Diphosphate as Adjunctive Therapy for Patients Hospitalized With Severe Acute Respiratory Syndrome Coronavirus 2 (SARS-CoV-2) Infection: A Randomized Clinical Trial. JAMA Netw Open. 2020 01;3(4):e208857.

93. Cantini F, Niccoli L, Matarrese D, Nicastri E, Stobbione P, Goletti D. Baricitinib therapy in COVID-19: A pilot study on safety and clinical impact. J Infect. 2020 Apr 23;

94. Hung IF-N, Lung K-C, Tso EY-K, Liu R, Chung TW-H, Chu M-Y, et al. Triple combination of interferon beta-1b, lopinavir-ritonavir, and ribavirin in the treatment of patients admitted to hospital with COVID-19: an open-label, randomised, phase 2 trial. Lancet Lond Engl. 2020 May 8;

95. Sanders JM, Monogue ML, Jodlowski TZ, Cutrell JB. Pharmacologic Treatments for Coronavirus Disease 2019 (COVID-19): A Review. JAMA. 2020 Apr 13;

96. Mehra MR, Desai SS, Ruschitzka F, Patel AN. Hydroxychloroquine or chloroquine with or without a macrolide for treatment of COVID-19: a multinational registry analysis. The Lancet [Internet]. 2020 May 22 [cited 2020 May 23];0(0). Available from: https://www.thelancet.com/journals/lancet/article/PIIS0140-6736(20)31180-6/abstract

97. Ren L, Xu W, Overton JL, Yu S, Chiamvimonvat N, Thai PN. Assessment of Hydroxychloroquine and Chloroquine Safety Profiles: A Systematic Review and Meta-Analysis. medRxiv [Internet]. 2020 May 8 [cited 2020 May 20];2020.05.02.20088872. Available from: https://www.medrxiv.org/content/10.1101/2020.05.02.20088872vl

98. Wang J, Yu L, Li K. Benefits and Risks of Chloroquine and Hydroxychloroquine in The Treatment of Viral Diseases: A Meta-Analysis of Placebo Randomized Controlled Trials. medRxiv [Internet]. 2020 Apr 18 [cited 2020 May 20];2020.04.13.20064295. Available from: https://www.medrxiv.org/content/10.1101/2020.04.13.20064295vl

99. Sarma P, Kaur H, Kumar H, Mahendru D, Avti P, Bhattacharyya A, et al. Virological and clinical cure in COVID-19 patients treated with hydroxychloroquine: A systematic review and meta-analysis. J Med Virol. 2020 Apr 16;

100. Yang T-H, Chou C-Y, Yang Y-F, Yang Y-P, Chien C-S, Yarmishyn AA, et al. Systematic Review and Meta-analysis of the Effectiveness and Safety of Hydroxychloroquine in COVID-19. medRxiv [Internet]. 2020 May 14 [cited 2020 May 20];2020.05.07.20093831. Available from: https://www.medrxiv.org/content/10.1101/2020.05.07.20093831v2

101. Yang Z, Liu J, Zhou Y, Zhao X, Zhao Q, Liu J. The effect of corticosteroid treatment on patients with coronavirus infection: a systematic review and meta-analysis. J Infect. 2020 Apr 10;

102. Gangopadhyay KK, Mukherjee JJ, Sinha B, Ghosal S. The role of corticosteroids in the management of critically ill patients with coronavirus disease 2019 (COVID-19): A meta-analysis. medRxiv [Internet]. 2020 Apr 22 [cited 2020 May 20];2020.04.17.20069773. Available from: https://www.medrxiv.org/content/10.1101/2020.04.17.20069773vl

103. Li H, Chen C, Hu F, Wang J, Zhao Q, Gale RP, et al. Impact of corticosteroid therapy on outcomes of persons with SARS-CoV-2, SARS-CoV, or MERS-CoV infection: a systematic review and meta-analysis. Leukemia. 2020 May 5;

104. Lu S, Zhou Q, Huang L, Shi Q, Zhao S, Wang Z, et al. Effectiveness and Safety of Glucocorticoids to Treat COVID-19: A Rapid Review and Meta-Analysis. medRxiv [Internet]. 2020 Apr 22 [cited 2020 May 20];2020.04.17.20064469. Available from: https://www.medrxiv.org/content/10.1101/2020.04.17.20064469vl

105. Shen C, Wang Z, Zhao F, Yang Y, Li J, Yuan J, et al. Treatment of 5 Critically Ill Patients With COVID-19 With Convalescent Plasma. JAMA. 2020 Mar 27;

106. Ahn JY, Sohn Y, Lee SH, Cho Y, Hyun JH, Baek YJ, et al. Use of Convalescent Plasma Therapy in Two COVID-19 Patients with Acute Respiratory Distress Syndrome in Korea. J Korean Med Sci. 2020 Apr 13;35(14):e149.

107. Rajendran K, Krishnasamy N, Rangarajan J, Rathinam J, Natarajan M, Ramachandran A. Convalescent plasma transfusion for the treatment of COVID-19: Systematic review. J Med Virol. 2020 May 1;

